# Unraveling the dynamics of the Omicron and Delta variants of the 2019 coronavirus in the presence of vaccination, mask usage, and antiviral treatment

**DOI:** 10.1101/2022.02.23.22271394

**Authors:** Calistus N. Ngonghala, Hemaho B. Taboe, Salman Safdar, Abba B. Gumel

## Abstract

The effectiveness of control interventions against COVID-19 is threatened by the emergence of SARS-CoV-2 variants of concern. We present a mathematical model for studying the transmission dynamics of two of these variants (Delta and Omicron) in the United States, in the presence of vaccination, treatment of individuals with clinical symptoms of the disease and the use of face masks. The model is parameterized and cross-validated using observed daily case data for COVID-19 in the United States for the period from November 2021 (when Omicron first emerged) to March 2022. Rigorous qualitative analysis of the model shows that the disease-free equilibrium of the model is locally-asymptotically stable when the control reproduction number of the model (denoted by ℝ_*c*_) is less than one. This equilibrium is shown to be globally-asymptotically stable for a special case of the model, where disease-induced mortality is negligible and both vaccine-derived immunity in fully-vaccinated individuals and natural immunity do not wane, when the associated reproduction number is less than one. The epidemiological implication of the latter result is that the combined vaccination-boosting strategy can lead to the elimination of the pandemic if its implementation can bring (and maintain) the associated reproduction number to a value less than one. An analytical expression for the vaccine-derived herd immunity threshold is derived. Using this expression, together with the baseline values of the parameters of the parameterized model, we showed that the vaccine-derived herd immunity can be achieved in the United States (so that the pandemic will be eliminated) if at least 68% of the population is fully-vaccinated with two of the three vaccines approved for use in the United States (Pfizer or Moderna vaccine). Furthermore, this study showed (as of the time of writing in March 2022) that the control reproduction number of the Omicron variant was approximately 3.5 times that of the Delta variant (the reproduction of the latter is computed to be ≈ 0.2782), indicating that Delta had practically died out and that Omicron has competitively-excluded Delta (to become the predominant variant in the United States). Based on our analysis and parameterization at the time of writing of this paper (March 2022), our study suggests that SARS-CoV-2 elimination is feasible by June 2022 if the current baseline level of the coverage of fully-vaccinated individuals is increased by about 20%. The prospect of pandemic elimination is significantly improved if vaccination is combined with a face mask strategy that prioritizes moderately effective and high-quality masks. Having a high percentage of the populace wearing the moderately-effective surgical mask is more beneficial to the community than having low percentage of the populace wearing the highly-effective N95 masks. We showed that waning natural and vaccine-derived immunity (if considered individually) offer marginal impact on disease burden, except for the case when they wane at a much faster rate (e.g., within three months), in comparison to the baseline (estimated to be within 9 months to a year). Treatment of symptomatic individuals has marginal effect in reducing daily cases of SARS-CoV-2, in comparison to the baseline, but it has significant impact in reducing daily hospitalizations. Furthermore, while treatment significantly reduces daily hospitalizations (and, consequently, deaths), the prospects of COVID-19 elimination in the United States are significantly enhanced if investments in control resources are focused on mask usage and vaccination rather than on treatment.

## 1. Introduction

The 2019 novel coronavirus (COVID-19), caused by SARS-CoV-2, resulted in an unprecedented pandemic never before seen since the 1918 influenza pandemic [1–5]. As of February 22, 2022, the SARS-CoV-2 pandemic had caused over 427.8 million confirmed cases and 5.9 million deaths globally [6, 7]. The rapid development and deployment of effective vaccines has played a major role in minimizing and mitigating the burden of the pandemic in regions with moderate and high vaccination coverage [8–10]. Specifically, as of February 11, 2022, 27 vaccines had been approved for use in numerous countries around the world [11]. Three of these vaccines, developed by Pfizer/BioNTech, Moderna and Johnson & Johnson, have been approved by the United States Food and Drugs Administration (FDA) [12, 13]. The Pfizer and Moderna vaccines are designed by introducing messenger ribonucleic acid (mRNA) that encodes the spike protein of SARS-CoV-2 to elicit an adaptive immune response against the disease [14]. These vaccines, which are primarily administered in a two-dose regimen three to four weeks apart, have an estimated protective efficacy against symptomatic COVID-19 infection of 95% [15–17]. A third (booster) dose was approved for these vaccines on November 19, 2021 for all adults from 18 years old [18]. The Johnson and Johnson vaccine, on the other hand, was developed using adenovirus vector encoding the SARS-CoV-2 spike protein, and is administered in a single dose regimen. Its efficacy against moderate to severe infection is estimated to be 67% [19]. We focus on two of the three vaccines used in the United States.

Despite the deployment of the three safe and effective vaccines in the United States (since late 2020 to early 2021), SARS-CoV-2 transmission in the United States is still on the rise (see, for example, Figure 1). Specifically, as of the time of writing (mid-February 2022), the United States is recording a 7-days average of 172,951 new COVID-19 cases *per* day [6]. This is largely due to vaccine hesitancy or refusal (only about 64% of the population in the United States is fully-vaccinated; with 43% of this population having received the booster dose as of February 14, 2022) and the emergence of new SARS-CoV-2 variants [20–25]. Thus, the approved vaccines, despite their high efficacy against the original SARS-CoV-2 strain, are currently insufficient to halt the COVID-19 pandemic in the United States. Hence, other control measures, such as antivirals that reduce the risk of disease progression, are needed to add to the armoury of measures for effectively combating or eliminating the COVID-19 pandemic in the United States. It is worth stating that the phenomenon of vaccine hesitancy or refusal during outbreaks of highly contagious and/or fatal vaccine-preventable infectious diseases of humans has a long history, dating back to the 1800s [26–30]. That is, vaccine hesitancy, refusal or general controversy surrounding vaccination programs in humans did not just start with the COVID-19 pandemic.

**Fig. 1:**
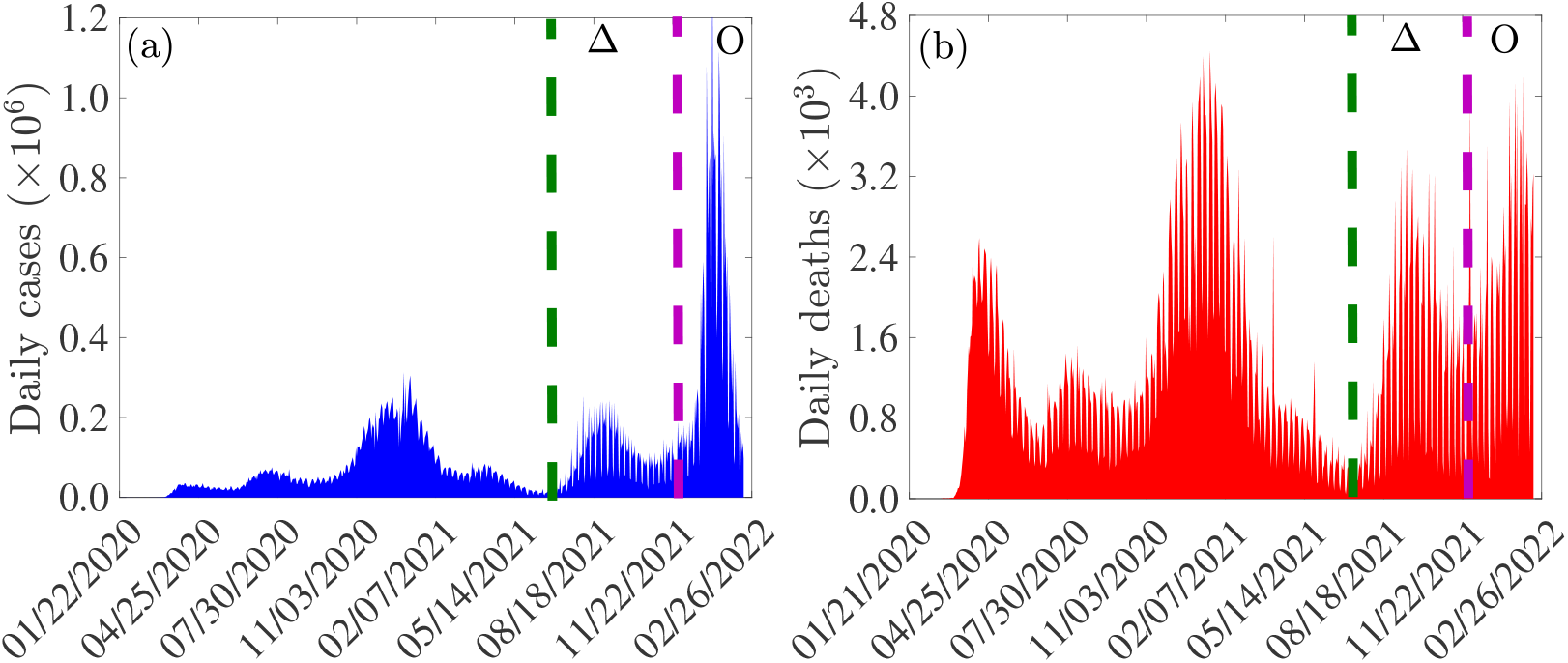
Data for COVID-19 daily (a) cases and (b) mortality for the United States for the period of the COVID-19 pandemic from January 2020 to February 2022. The data is obtained from the Johns Hopkins University COVID-19 Dashboard [7, 44]. The burden of the Delta variant is in the regions between the green and purple dashed vertical lines (denoted by Δ), while that of the Omicron variant is in the region to the right of the dashed purple vertical line (denoted by *O*).

Two antiviral drugs, namely, *Paxlovid* (developed by Pfizer Inc.) and *Molnupiravir* (developed by Merck Inc.), received FDA-Emergency Use Authorization in December 2021, for healthcare providers to use to treat mild-to-moderate COVID-19 in adults and pediatric patients (12 years of age and older weighing at least 40 kg) [31]. These antivirals primarily work by altering the genetic code of SARS-CoV-2 and inhibiting it from replicating. Paxlovid, which is administered as three tablets taken together orally twice daily for five days, is estimated to reduce the risk of hospitalization or death by 90% [32]. Molnupiravir, which is administered as four 200 milligram capsules taken orally every 12 hours for five days, is effective in reducing hospitalization or death by 31% [33]. Full efficacy of these antivirals is achieved if used during the first five days of onset of symptoms [33, 34].

The unprecedented level of community transmission of SARS-CoV-2 in the United States has resulted in the emergence of numerous mutants (variants of concern) [35–40]. The Delta variant (B.1.617.2), which was first identified in India in late 2020 [41], emerged in the United States in July 27, 2021. This variant, which was believed to be more than twice as contagious as previous SARS-CoV-2 variants, quickly spread across the United States causing unprecedented levels of hospitalizations and deaths. It remained the dominant variant (accounting for 99% of all new cases) until mid-December 2021 (see the region between the dashed green and purple vertical lines in Figure 1), when a new variant (Omicron; B.1.1.529) emerged [37]. Omicron, believed to be three times more contagious than Delta, quickly displaced Delta, and became the predominant variant [17, 42, 43] (see regions to the right of the dashed purple vertical line in Figure 1).

The three FDA-approved vaccines used in the United States were designed against the original SARS-Cov-2 strain, and only offer cross-protective efficacy against the variants [14, 17, 45]. It is, therefore, instructive to theoretically assess, through mathematical modeling and analysis, the population-level effectiveness of the vaccines against the two remaining dominant variants (namely, Delta and the current predominant, Omicron). The current study is based on the design of a mathematical model for assessing the qualitative dynamics of the two dominant SARS-CoV-2 variants co-circulating in the United States (Delta and Omicron). The resulting two-strain model, which takes the form of a deterministic system of nonlinear differential equations, incorporates treatment of individuals with clinical symptoms of SARS-CoV-2 (using the two FDA-EUA antivirals discussed above). The model is formulated in Section 2.1 and fitted using daily case data for COVID-19 in the United States for the period when Omicron first appeared in the country (i.e., from November 28, 2021 to January 31, 2022), as well as cross-validated using the portion of the daily case data from February 1, 2022 to March 18, 2022 and cumulative case data for COVID-19 in the United States (for the period from November 28, 2021 to March 18, 2022) in Section 2.2. Basic theoretical results, for the existence and asymptotic stability of the disease-free equilibrium of the model, are provided in Section 3. Furthermore, the condition, in parameter space, for achieving vaccine-derived herd immunity is derived. Numerical simulations are presented in Section 4 and a discussion of the results and concluding remarks are presented in Section 5. It should be mentioned that this study was conducted between November 2021 to March 2022 (and the reader should interpret our results bearing this context and fact in mind).

## 2. Methods

### 2.1. Formulation of Mathematical Model

The mathematical model to be developed in this study is for the transmission dynamics of the COVID-19 pandemic in the United States for the scenario where the dominant variant of concern was initially Delta and the new prevailing and currently dominant variant, Omicron, is introduced (this was the scenario in the United States when Omicron emerged in the Fall of 2021). Let *N*(*t*) denote the total population of the United States at time *t*. This population is stratified into the mutually-exclusive compartments for unvaccinated susceptible (*S*(*t*)), fully-vaccinated but not boosted susceptible (*V*_*f*_(*t*); these are individuals that have either received the two doses of the FDA-approved Pfizer or Moderna vaccine or the single dose of Johnson & Johnson vaccine), fully-vaccinated and boosted susceptible (*V*_*b*_(*t*)), exposed/latent (*E*_*j*_ (*t*)), presymptomatic (*P*_*j*_(*t*)), detected infected individuals from the exposed, presymptomatic, and asymptomatic classes (*Q*_*j*_(*t*)), symptomatic infectious within the first five days of onset of symptoms (*I* _*j*1_(*t*)), symptomatic individuals after the first five days of onset of symptoms (*I* _*j*2_(*t*)), asymptomatic infectious or symptomatic infectious with very mild symptoms (*A*_*j*_(*t*)), hospitalized (*H*_*j*_(*t*)) and recovered (*R*_*j*_(*t*)) individuals, so that (where we used the subscript notation *j* ∈ {*d,o*}, with *d* and *o* representing the Delta and Omicron variant, respectively):

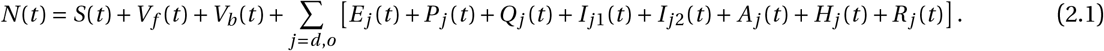

The population of unvaccinated susceptible individuals is increased by recruitment into the population at a rate Λ and by the waning of vaccine-derived protective immunity in fully-vaccinated but not boosted (at a rate *ω*_*v f*_) and fully-vaccinated and boosted (at a rate *ω*_*vb*_) individuals. It is further increased by the loss of natural immunity in individuals who recovered from Delta (at a rate *ω*_*dr*_) and Omicron (at a rate *ω*_*or*_) infection. Unvaccinated susceptible individuals are fully-vaccinated (but not boosted) at a rate *ξ*_*v f*_. Individuals in all epidemiological compartments suffer natural death at a rate *µ*. Unvaccinated susceptible individuals acquire infection with the Delta variant at a rate *λ*_*d*_ and with Omicron at a rate *λ*_*o*_, where:

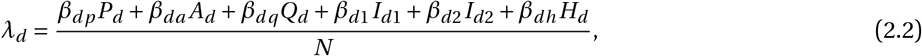

and,

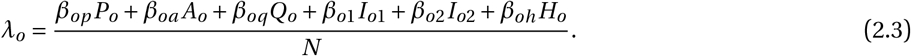

In (2.2)−(2.3), *β*_*jp*_, *β*_*ja*_, *β*_*jq*_, *β*_*ji*_ and *β*_*jh*_ (with *j* = *d, o, i* = 1,2) represent, respectively, the transmission rate by presymptomatic, asymptomatic, detected, symptomatic, and hospitalized infectious individuals. It should be mentioned that detected individuals are supposed to be isolated (or removed) from the actively-mixing population. However, in reality, isolation is not always perfectly implemented (leading to the well-known notion of *leaky isolation*, where some isolated individuals escape isolation for all sorts of reasons, such as economics (to allow them go to work; particularly those who live from paycheck-to-paycheck) or human behavior change, leading them to choose to opt out of the isolation protocols). Our modeling allows for the possibility that not all detected individuals will strictly adhere to the isolation protocols (so that some level of disease transmission can occur by detected individuals who are supposed to be “isolated”). This is reflected in our parameter estimation in Section 2.2, where the estimated value for the transmission rates for detected individuals are very small, compared to those for infectious individuals who are not in isolation (see Table S4 of the Supplementary Information). Furthermore, it should be noted that individuals entering the *Q*_*j*_ class from the *E* class are infected but not (yet) infectious. In other words, not all individuals in the *Q*_*j*_ class are infectious. We account for this fact by re-defining the effective contact rate for individuals in the *Q*_*j*_ class (*β*_*jq*_, *j* = *d,o*) as 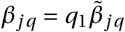, where 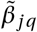 is the overall effective contact rate for individuals in the *Q*_*j*_ class and 0 < *q*_1_ ≤ 1 is a modification parameter accounting for the proportion of individuals in the *Q*_*j*_ class who are actually infectious.

Fully-vaccinated (but not boosted) individuals acquire breakthrough infection with Delta (at a rate *λ*_*d f*_) or Omicron (at a rate *λ*_*o f*_), where:

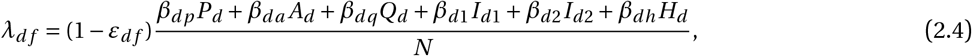

and,

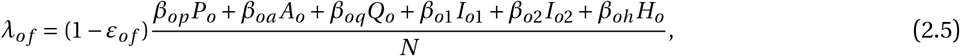

with 0 < *ε*_*j f*_ < 1 representing the cross-protective efficacy the vaccine offers against Delta (*ε*_*d f*_) or Omicron (*ε*_*o f*_) variant. Fully-vaccinated individuals receive the booster dose (at a rate rate *ξ*_*vb*_) and lose their vaccine-derived immunity (at a rate *ω*_*v f*_). Vaccinated individuals who received the booster dose acquire infection with Delta (at a rate *λ*_*db*_) or Omicron (at a rate *λ*_*ob*_), where:

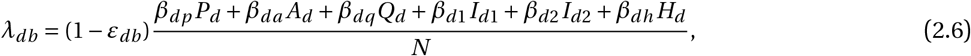

and,

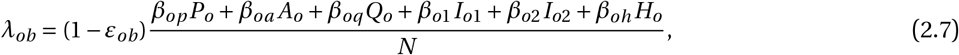

with 0 < *ε*_*db*_ < 1 and 0 < *ε*_*ob*_ < 1 representing the cross-protective efficacy the vaccine offers boosted individuals against Delta and Omicron, respectively. Exposed (or latent) individuals progress to the pre-symptomatic class at a rate *σ*_*je*_. Furthermore, individuals in the pre-symptomatic class progress, at the end of the incubation period, to either the symptomatic class (at a rate *r* _*j*_ *σ*_*jp*_; where 0 < *r* _*j*_ < 1 is the proportion of these individuals that display clinical symptoms of SARS-CoV-2 at the end of the incubation period) or the asymptomatic infectious class (at a rate (1 − *r* _*j*_)*σ*_*jp*_). Asymptomatic infectious individuals recover (naturally) at a rate *γ*_*ja*_. Infected individuals in the exposed, presymptomatic and asymptomatic classes are detected at rate *ρ*_*j*_, detected cases become symptomatic at rate *ψ*_*j*_, recover naturally at rate *γ*_*jq*_, or die naturally at rate *µ*. Symptomatic individuals during the first five days of onset of symptoms are treated at a rate *τ*_*j*1_ and progress to the second infectious class at a rate *α*_*j*1_. These individuals are hospitalized at a rate *ϕ*_*j*1_ and suffer disease-induced death at a rate *δ*_*j*1_. Symptomatic individuals that survived the first five days of onset of symptoms are treated at a rate *τ*_*j* 2_, hospitalized at a rate *ϕ*_*j*2_, recover at a rate *γ*_*j*2_ and die of the disease at a rate *δ*_*j*2_. Hospitalized individuals are treated at a rate *τ*_*jh*_ and succumb to the disease at a rate *δ*_*jh*_. Recovered individuals lose the infection-acquired (natural) immunity, and revert to the wholly-susceptible class, at a rate *ω*_*jr*_.

The equations for the transmission dynamics of the two variants of concern (Delta and Omicron) in the United States, in the presence of vaccination with the FDA-approved Pfizer/BioNTech or Moderna vaccine and treatment of symptomatic individuals, is given by the following deterministic system of nonlinear differential equations (the flow diagram of the model is given in Figure 2 and the state variables and parameters of the model are described in Tables S1 and S2 of the Supplementary Information, respectively):

**Fig. 2:**
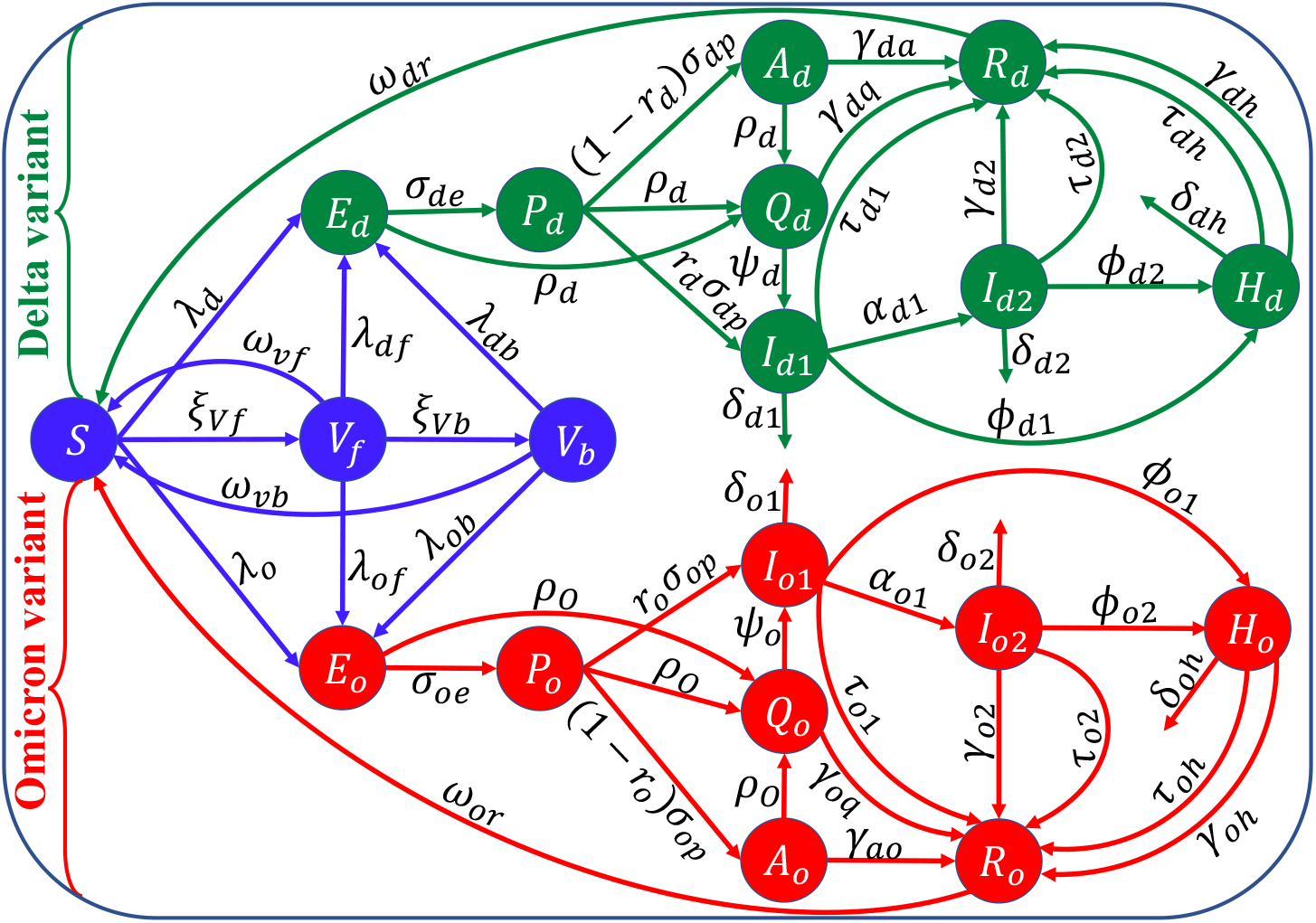
Flow diagram of the model (2.8). Although recruitment into the population and natural deaths occur (at the rate Λ and *µ*, respectively), these rates are not illustrated in the flow diagram to make it less crowded and easier to follow. The state variables and parameters are described in Tables S1 and S2 of the Supplementary Information (SI).

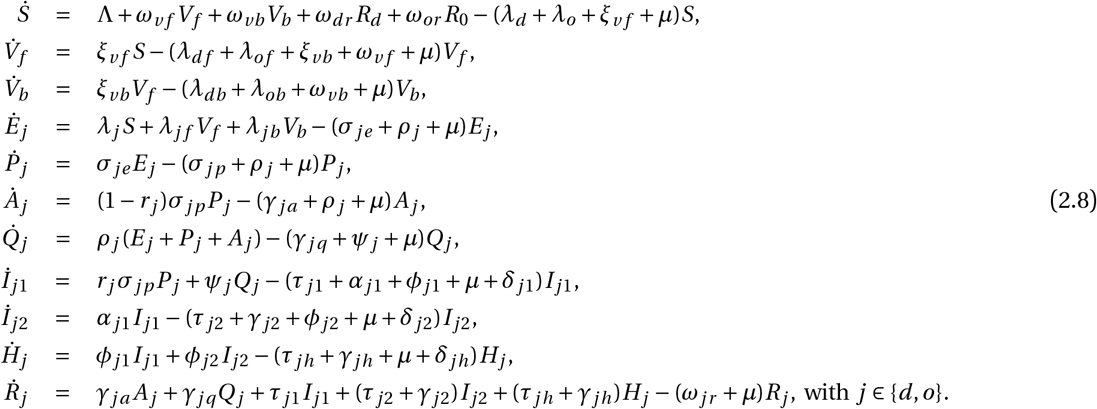

Some of the main assumptions made in the formulation of the model (2.8) are

a. Only the Delta and Omicron variants were co-circulating (i.e., they were the most predominant of all the variants of concern at the time of this study). Specifically, we consider the scenario where Delta was the dominant strain and then Omicron emerged (as was the case in the United States during the fall of 2021).
b. A well-mixed population, where every individual is equally likely to mix with every other individual in the community. We also consider an endemic scenario, to account for demographic (birth and natural death) processes. This is needed to account for the vaccination of children (five years of age and older).
c. Vaccines only offer cross-protective efficacy against the variants (since all the FDA-approved vaccines were developed for the original strain that was circulating in the United States during the early stages of the pandemic).
d. Natural immunity and vaccine-derived immunity for fully-vaccinated and boosted humans wane over time.

The model (2.8) is an extended version of numerous COVID-19 transmission models in the literature that consider the dynamics of multiple co-circulating variants, such as those in [46–55] by, *inter alia*, including:

1. Incorporating a booster dose (this was not considered in [46–55]).
2. Explicitly modeling the dynamics of the two SARS-CoV-2 variants (Delta and Omicron) that were predominant at the time this study was carried out and submitted (November 2021 to March 2022).
3. Incorporating the treatment of symptomatic individuals during and after the first five days of onset of symptoms (treatment against COVID-19 was not considered in [46–54]). It is worth mentioning that although the agent-based model considered in Matrajt *et al*. [55] was used to evaluate the impact of treatment on COVID-19 dynamics, the model does not explicitly account for waning of immunity and the use of a booster shot against the disease.
4. Allowing for waning vaccine-derived and natural immunity over time (this was not considered in [46–55]).
5. Explicitly including the impact of voluntary testing to detect infected individuals who do not have clinical symptoms of the disease. Although this was not done in [46–55], detection has been incorporated in some models for COVID-19 dynamics (e.g., [56, 57]). Specifically, the epidemic (SIDARTHE) model developed in [56] consists of eight epidemiological compartments of Susceptible (*S*), Infected (*I*), Diagnosed (*D*), Ailing (*A*), Recognized (*R*), threatened (*T*), Healed (*H*) and Extinct (*E*) individuals, where (in the formulation/notation in [56]) individuals in the *I* and *A* compartments are undetected while those in the *D, R*, and *T* are detected. Important differences exist between the epidemic SIDARTHE model formulated in [56] and the endemic SVEPAIQHR (Susceptible-Vaccinated-Exposed-Presymptomatic-Asymptomatic-Symptomatic-Detected-Hospitalized-Recovered) model presented in the current study. For instance, the model in [56] did not explicitly include vaccination, births and natural deaths. Furthermore, it assumes that newly-infected individuals are instantaneously capable of transmitting infection (i.e., susceptible individuals move to the *I* class upon acquisition of infection; this may not be realistic in the context of COVID-19, where infected individuals must first survive either latency (for presymptomatic infectious individuals) or incubation period (for asymptomatic or symptomatic infectious individuals) before they can transmit infection (i.e., before they become infectious). It should also be stated that detection of infected individuals who do not display clinical symptoms of the disease (i.e., those in the exposed/latent class, *E*, presymptomatic class, *P*_*j*_, *j* = *d,o*, and asymptomatic class, *A*_*j*_) is crucial in allowing the model to fit the daily confirmed case data reasonably well (we would not get a good model fitting, with respect to the daily confirmed case data if this feature is not included in the model). In particular, including detection of infected individuals in these classes that do not exhibit disease symptoms makes it possible to be able to fit confirmed daily cases in the model to confirmed daily cases from the available data [56]. Finally, although voluntary testing is modeled in our study using constant parameters (*τ*_*ji*_ and *τ*_*jh*_, where *j* = *d,o*, and *i* = 1,2) (as a first approximation, necessitated by the absence of real data on human behavior with respect to testing in the United States), it may be more appropriate to use time-varying parameters that depend on human behavioral changes to model voluntary testing (as was done in some studies, such as those in [58, 59]).

### 2.2. Model Fitting and Parameter Estimation

The model (2.8) has several parameters, some of which are known (see Table S3) and some of which are unknown. The key unknown parameters that will be estimated are the effective community transmission rates for infectious individuals in the classes *P*_*d*_, *A*_*d*_, *Q*_*d*_, *I*_*d*1_, *I*_*d*2_, and *H*_*d*_ (i.e., *β*_*dp*_, *β*_*da*_, *β*_*dq*_, *β*_*d*1_, *β*_*d*2_, and *β*_*dh*_), the rate at which individuals are fully-vaccinated (*ξ*_*v f*_), and the rate at which fully-vaccinated individuals are boosted (*ξ*_*vb*_). These parameters will be estimated by fitting the model (2.8) to the confirmed daily COVID-19 case data for the United States for the period from November 28, 2021 to January 31, 2022. The model (2.8) is fitted to (and cross-validated) using the observed data using a standard nonlinear least squares regression model fitting approach [60–62]. Specifically, the fitting and parameter estimation involves computing the best set of parameter values that minimizes the sum of the square difference between the observed new daily confirmed case data and the new daily cases from the model (2.8) (i.e., *ρ*_*d*_ (*E*_*d*_ +*P*_*d*_ + *A*_*d*_)+*r*_*d*_*σ*_*dp*_*P*_*d*_+*ρ*_*o*_(*E*_*o*_ +*P*_*o*_ + *A*_*o*_)+*r*_*o*_*σ*_*op*_*P*_*o*_). The minimization is implemented in MATLAB version R2021b using the inbuilt “*lsqcurvefit*” algorithm. The 95% confidence intervals for the estimated parameters are determined using a bootstrapping technique [60–62]. Bootstrapping involves producing a large collection of simulated data sets from a given data set by sampling from this given data set with replacement and then using each generated data set to estimate model parameters. These estimations are used for setting confidence intervals based on the distribution of the estimates, which is associated with the actual estimates. In this study, we sample the residuals from the initial parameter estimation using the inbuilt bootstrapping function in MATLAB (“*bootstrp*”) to generate 10,000 bootstrap replicates. The bootstrap data sets are generated by adding the resampled residuals to the best fit curve. The model is then fitted to each bootstrap data set to create the bootstrap distribution of parameter estimates. This distribution is used to estimate the 95% confidence intervals of the parameters via the inbuilt function (“*prctile*”) in MATLAB. Fitting the model to (raw) daily confirmed case data is useful in avoiding mistakes that arise when deterministic models are fitted to cumulative case data [63]. The estimated parameters and the corresponding 95% confidence intervals are tabulated in Table S4.

The results obtained for the fitting of the daily new COVID-19 cases in the United States, for the period when Omicron first emerged (November 28, 2021) until January 31, 2022 (i.e., the region to the left of the dashed vertical black line), is depicted in Figure 3 (a). This figure shows a very good fit between the model output (blue curve) and the observed data (red dots). Furthermore, we show in this figure the prediction of the model for the daily COVID-19 cases for approximately a seven-week period after January 31, 2022 (i.e., for the period from February 1, 2022 to March 18, 2022), as illustrated by the segment of the graph to the right of the dashed vertical black line of Figure 3 (a). This segment of Figure 3 (a) clearly shows that the model (2.8) predicts the observed data for the period from February 1, 2022 to March 18, 2022 perfectly (solid green curve). The model was then simulated (using the fixed and fitted parameter baseline values in Tables S3 and S4) and compared with the observed cumulative case data. The results obtained, depicted in Figure 3 (b), show a very good fit. Furthermore, this figure shows a perfect model prediction for the period from February 1, 2022 to March 18, 2022 (see region to the right of the dashed vertical black line).

**Fig. 3:**
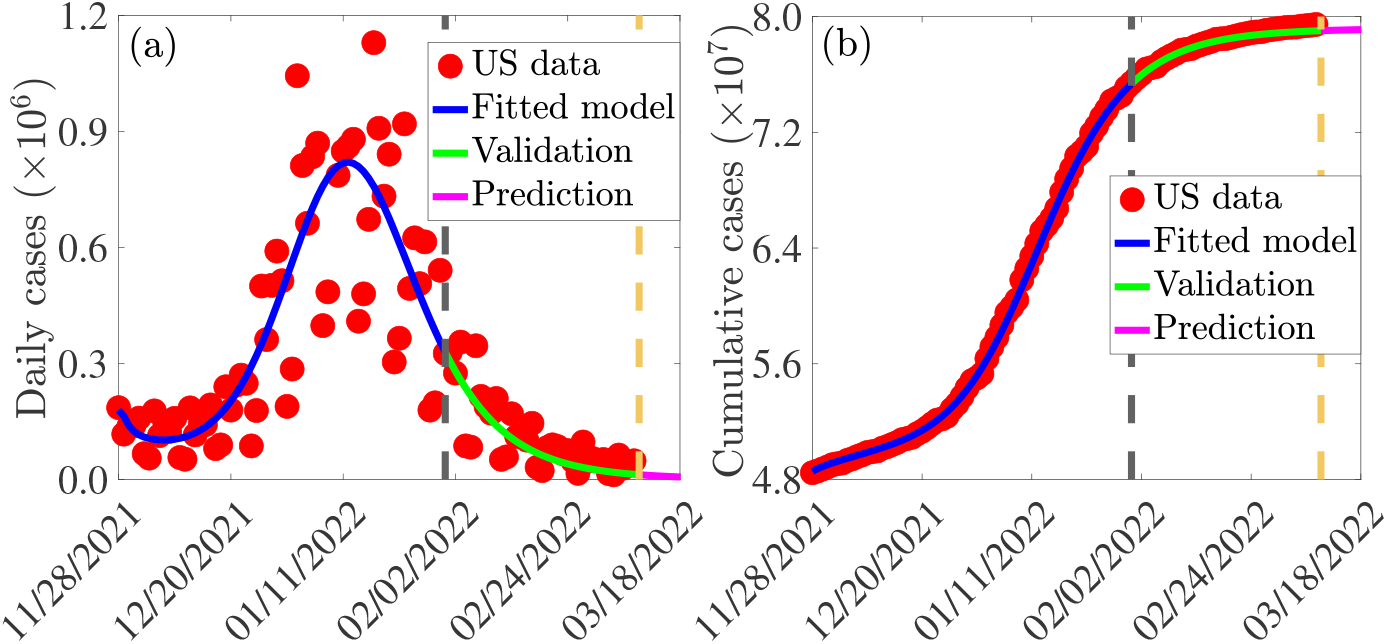
(a) Time series illustration of the least squares fit of the model (2.8), showing the model’s output for the daily cases (blue curve) compared to the observed daily confirmed cases for the United States (red dots) from November 28, 2021 to January 31, 2022 (segment to the left of the dashed vertical black line). (b) Simulation result of the model (2.8), showing cumulative COVID-19 cases for the United States as a function of time, using the fixed and estimated baseline parameter values given in Tables S3 and S4. The segment from February 1, 2022 to March 18, 2022 (i.e., solid green and magenta curves or the entire segment to the right of the dashed black vertical line) illustrates the performance of the model (2.8) in predicting the daily and cumulative cases in the United States.

## 3. Theoretical Analysis

In this section, qualitative properties of the model (2.8), with respect to the disease-free equilibrium (DFE), will be explored. The DFE of the model (2.8) is given by:

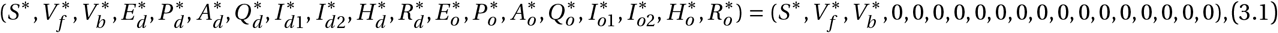

where,

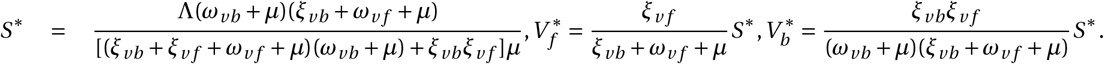

### 3.1. Asymptotic Stability of DFE

#### 3.1.1. Local asymptotic stability of DFE

The *vaccination reproduction number* of the model, with respect to variant *j* (with *j* ∈ {*d,o*}), denoted by ℝ_*jv*_, can be obtained using the *next generation operator method* [64, 65]. It is given by the following expression:

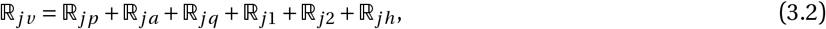

where,

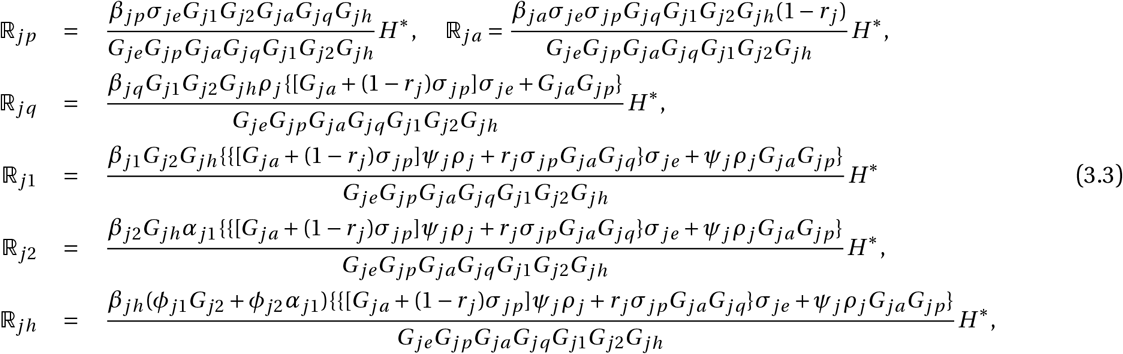

with,

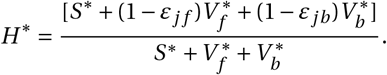

In (3.3), *G*_*je*_ = *σ*_*je*_ + *ρ*_*j*_ + *µ,G*_*jp*_ = *σ*_*jp*_ + *ρ*_*j*_ + *µ,G*_*ja*_ = *γ*_*ja*_ + *ρ*_*j*_ + *µ,G*_*jq*_ = *γ*_*jq*_ + *ψ*_*j*_ + *µ*;*G*_*j*1_ = *τ*_*j*1_ + *α*_*j*1_ + *ϕ*_*j*1_ + *µ* + *δ*_*j*1_,*G*_*j*2_ = *τ*_*j*2_ + *γ*_*j*2_ + *ϕ*_*j*2_ + *µ* + *δ*_*j*2_ and *G*_*jh*_ = *γ*_*jh*_ + *τ*_*jh*_ + *µ* + *δ*_*jh*_. It is convenient to define:

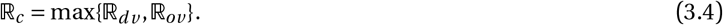

The quantity ℝ_*c*_ is the *vaccination reproduction number* of the model (2.8). It measures the average number of new SARS-CoV-2 cases generated by a typical infectious individual introduced in a community where a certain proportion of the susceptible individuals is fully-vaccinated. The result below follows from Theorem 2 of [65].

##### Theorem 3.1.

*The DFE of the model* (2.8) *is locally-asymptotically stable if* ℝ_*c*_ < 1, *and unstable if* ℝ_*c*_ > 1.

The epidemiological implication of Theorem 3.1 is that a small influx of infected individuals will not generate a large outbreak of the SARS-CoV-2 virus in the community. Thus, the SARS-CoV-2 pandemic can be controlled effectively if the control reproduction number of the model (ℝ_*c*_) can be brought to a value less than one (and maintained), provided the initial number of infected individuals is small enough. Mathematically-speaking, although bringing ℝ_*c*_ to a value less than one is necessary for elimination of the SARS-CoV-2 virus, it may not be sufficient (owing to the possible existence of a stable endemic equilibrium when ℝ_*c*_ < 1, which is well-known to occur for disease transmission models with imperfect vaccines [66–68]). For effective control or elimination of the SARS-CoV-2 virus pandemic to be independent of the initial sizes of the sub-populations of the model, it is crucial to show that the disease-free equilibrium of the model (2.8) is globally-asymptotically stable. This is done, for a special case of the model, in Section 3.1.2 below.

#### 3.1.2. Global asymptotic stability of DFE: special case

Here, the global asymptotic stability of the disease-free equilibrium of the model will be explored for a special case. Consider a special case of the model (2.8) with negligible disease-induced mortality (i.e., *δ*_*j*1_ = *δ*_*j*2_ = *δ*_*jh*_ = 0, for *j* ∈ {*d,o*}) and no waning of vaccine-derived protective immunity for fully-vaccinated (but not boosted) individuals (i.e., *ω*_*v f*_ = 0) and no waning of natural immunity (i.e., *ω*_*dr*_ = *ω*_*or*_ = 0). Assuming negligible disease-induced mortality is reasonable since at the time of writing, the Omicron variant (which has been the dominant SARS-CoV-2 variant circulating in the United States since its emergence in the fall of 2021), although highly transmissible, was very marginally fatal, in comparison to Delta or any of the other SARS-CoV-2 variants that emerged in the United States [69–72]. The assumptions on waning immunity are made for mathematical tractability. Consider the following feasible region for the model (2.8):

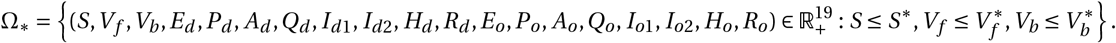

It can be shown (see Section S3.1 of the SI for the proof) that the region Ω_*_ is positively-invariant and attracting with respect to the special case of the model (2.8). Furthermore, it is convenient to define the following threshold quantity:

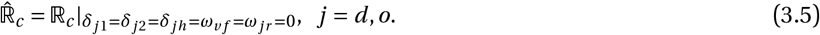

We claim the following result:

##### Theorem 3.2.

*Consider the special case of the model* (2.8) *in the absence of disease-induced mortality (i.e*., *δ*_*j* 1_ = *δ*_*j* 2_ = *δ*_*jh*_ = 0*) and no waning of vaccine-derived immunity in fully-vaccinated individuals (i.e*., *ω*_*v f*_ = 0*) and no waning of natural immunity (i.e*., *ω*_*dr*_ = *ω*_*or*_ = 0*). The disease-free equilibrium of the special case of the model (given in Eq*. (3.1) *with ω*_*v f*_ = 0*) is globally-asymptotically stable in* Ω_*_ *whenever* 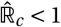.

The proof of Theorem 3.2, based on using comparison theorem [73], is given in the SI (Section S3.2). The epidemiological implication of Theorem 3.2 is that, for the special case of the model considered above, the COVID-19 pandemic can be effectively eliminated in the United States if the vaccination strategy implemented can result in bringing (and maintaining) the associated control reproduction number 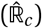 to a value less than one. Mathematically-speaking, Theorem 3.2 implies that 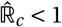 is necessary and sufficient for the elimination of the pandemic in the United States.

It is worth mentioning that, using the baseline values of the fixed and estimated parameters of the model (2.8), given in Tables S3 and S4 (used to generate the fittings in Figure 3), shows that the constituent reproduction numbers for the Delta (ℝ_*dv*_) and Omicron (ℝ_*ov*_) are ℝ_*dv*_ = 0.2782 < 1 (with 95% confidence interval ℝ_*dv*_ ∈ (0.1991, 0.5197)) and ℝ_*ov*_ = 0.9602 (with 95% confidence interval, ℝ_*ov*_ ∈ (0.6206, 1.7509)), so that the overall vaccination reproduction number of the model (ℝ_*c*_) = max{ℝ_*dv*_, ℝ_*ov*_} = ℝ_*ov*_ = 0.9602. Similarly, for the aforementioned special case of the model, the associated control reproduction numbers for the Delta and Omicron variants are given, respectively, by 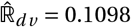 (with 95% confidence interval 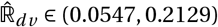) and 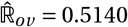 (with 95% confidence interval, 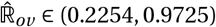). Hence, for the special case of the model, the overall vaccination reproduction number 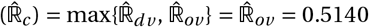. This suggests that, based on the data for the COVID-19 dynamics in the United States for the period November 28, 2021 to January 31, 2022, the Delta variant has essentially died out (owing to the very low value of its associated control reproduction number 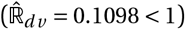) and that the Omicron variant has displaced the Delta variant (since its control reproduction number is larger) and become the predominant variant in the United States. Furthermore, since the reproduction number of Omicron (and, hence, of the model (2.8) itself) is less than one, it follows (from Theorem 3.2 or, equivalently, Theorem 3.3 below) that the Omicron variant may also be dying out if the baseline levels of the COVID-19 control measures being implemented in the United States (as of March 2022) are maintained. In other words, our analysis and data fitting, using data (as of March 2022) suggests that the COVID-19 pandemic in the United States may be entering its declining phase, and that elimination is feasible if baseline levels of the control measures being implemented are maintained.

### 3.2. Computation of Vaccine-derived Herd Immunity Threshold

To obtain an expression for the vaccine-derived herd immunity threshold [54, 68, 74], it is instructive to recall that the basic reproduction number for variant *j*, denoted by ℝ_0*j*_, with *j* ∈ {*d,o*}, is obtained from (3.2) by setting *V*_*f*_ = *V*_*b*_ = *ε*_*j f*_ = *ε*_*jb*_ = 0. It follows that ℝ_0 *j*_ = ℝ_*c*_|*V*_*f*_ =*V*_*b*_ =*ε* _*j f*_ =*ε*_*jb*_ =0. Let *f*_*v*_ = min{*f*_*v f*_, *f*_*vb*_} be the proportion of individuals who are fully-vaccinated only (*f*_*v f*_) and those who are fully-vaccinated and boosted (*f*_*vb*_) at steady-state. Furthermore, let *ε*_*jv*_ = min{*ε*_*j f*_, *ε*_*jb*_} be the minimum vaccine efficacy for fully-vaccinated (*ε*_*j f*_) and boosted (*ε*_*jb*_) individuals. Setting ℝ_*c*_ = 1 in (3.4), and solving for *f*_*v*_, gives the following expression for the vaccine-derived herd immunity threshold for Model (2.8):

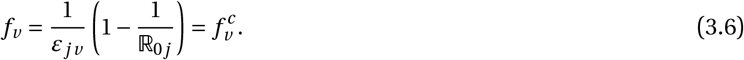

It follows from (3.6) that ℝ_*c*_ < (>)1 if 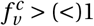. Thus, vaccine-induced herd immunity can be achieved in the community (and the disease can be eliminated) if 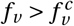. Theorem 3.1 can be written in terms of the herd immunity threshold as:

#### Theorem 3.3.

*The DFE of Model* (2.8) *is locally-asymptotically stable if* 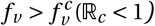, *and unstable if* 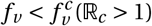.

Using the baseline parameter values in Tables S3 and S4, and the herd immunity threshold expression (3.6) shows that the value of the vaccine-derived herd immunity threshold for the United States is 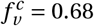. Thus, our study shows that vaccine-derived herd immunity can be achieved in the United States (during the time when both Omicron and Delta are circulating, with the former being the overwhelming predominant variant) if at least 68% of individuals living in the United States are fully-vaccinated with either the Pfizer and Moderna vaccine. In line with Theorem 3.3, COVID-19 can be eliminated in the United States if this level of vaccination coverage is attained. Data from [75, 76] shows that about 64% of the United States population is fully-vaccinated as of February 14, 2022. Hence, our study shows that the United States can eliminate the pandemic if approximately an additional 4% of unvaccinated individuals or individuals who have received only one dose of the Pfizer or Moderna vaccine become fully-vaccinated with either of these two vaccines.

## 4. Numerical Simulations

The model (2.8) is simulated, using the fixed and fitted baseline parameter values in Tables S3 and S4, to assess the impact of vaccination, boosting of vaccine-derived immunity, testing (and detection of infected individuals with no clinical symptoms of the disease) and treatment of symptomatic individuals on the dynamics of COVID-19 in the United States.

Figure 4 depicts contour plots of the control reproduction number of the model, as a function of the efficacy of the two vaccines (Pfizer or Moderna) against acquisition of infection with Delta or Omicron (defined as *ε*_*v*_ = min{*ε*_*v f*_, *ε*_*vb*_}) and the fraction of the United States population fully-vaccinated at steady-state (defined as *f*_*v*_ = min{ *f*_*vd*_, *f*_*vo*_}). For the case where mask usage in the community is maintained at baseline level, this figure shows a decrease in the reproduction number with increasing values of the vaccine efficacy and coverage. Specifically, for the case when the cross-protective vaccine efficacy against the two variants is set at 60%, population-wide vaccine-derived herd immunity (associated with the reduction of the reproduction number to a value below one) can be achieved if 84% of the United States population is fully-vaccinated with either the Pfizer or Moderna vaccine (Figure 4(a)). It follows from this contour plot that the herd immunity requirement reduces to 65% if the vaccines offer 80% cross-protective efficacy against the two variants.

**Fig. 4:**
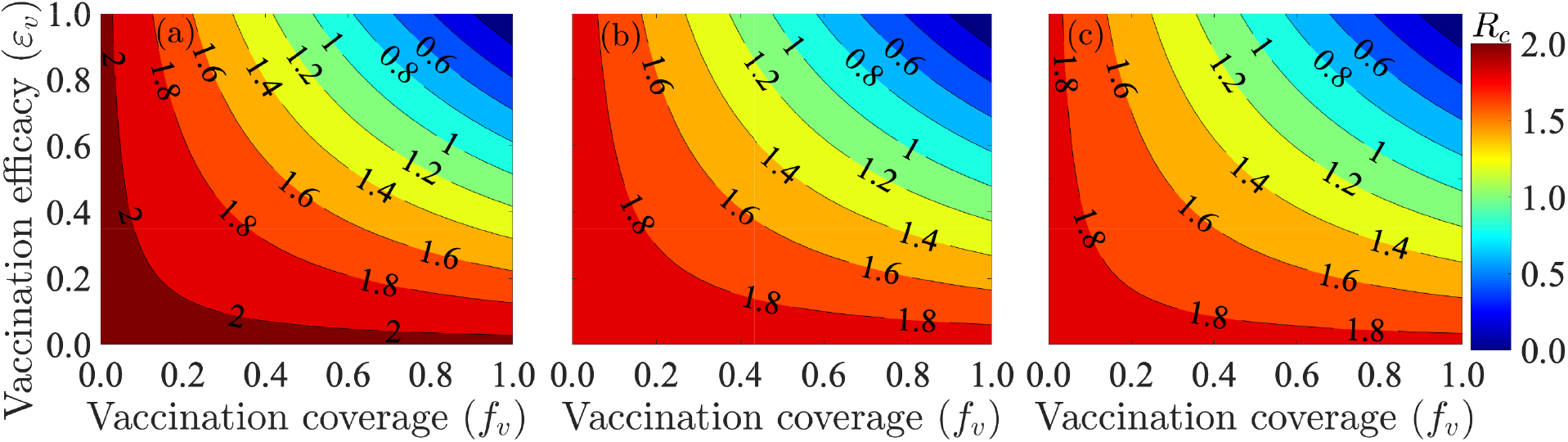
Contour plots of the control reproduction number of the model (2.8), ℝ_*c*_, as a function vaccine coverage (i.e., proportion of fully-vaccinated individuals, *f*_*v*_) and cross-protective vaccine efficacy against the variants (*ε*_*v*_ = min(*ε*_*v f*_, *ε*_*vb*_)) for the case when (a) mask coverage is maintained at its baseline value, (b) surgical mask is prioritized and the coverage in its usage is increased by 10% from its baseline value, (c) N95 mask is prioritized and the coverage in its usage is increased by 10% from its baseline value. The values of all other parameters used in the simulations are as given by the baseline values in Tables S3 and S4.

Further simulations are carried out for the case where the baseline face mask usage in the community is increased by 10%, for various mask types. Figure 4(b) shows that if surgical masks are prioritized, the herd immunity requirement corresponding to 60% (80%) cross-protective vaccine efficacy against the variants now reduces to 80% (60%), in comparison to the 84% (65%) coverage recorded for the case with baseline face mask coverage depicted in Figure 4(a). Furthermore, if N95 masks are prioritized, the herd immunity requirement corresponding to the cross-protective vaccine efficacy of 60% (80%) further reduces to 77% (58%). In summary, the contour plots in Figure 4(b) and (c) show that the proportion of individuals who need to be fully-vaccinated to achieve herd immunity reduces with increasing coverage of masks in the community (from the baseline mask usage), and the level of reduction achieved depends on the quality of the mask used (specifically, greater reduction in herd immunity level needed to eliminate the disease is achieved if the high-quality N95 masks are prioritized, in comparison to the scenario where the moderate quality surgical masks are prioritized).

### 4.1. Assessing the Impact of Vaccination Coverage

The impact of vaccination coverage (i.e., the rate at which unvaccinated susceptible individuals become fully-vaccinated) is monitored by simulating the model (2.8) with various values of the vaccination rate (*ξ*_*v f*_). To exclusively monitor the impact of vaccination, the simulations are carried out for the special case of the model with no treatment (i.e., all treatment-related parameters and state variables of the model are set to zero). The simulation results obtained, depicted in Figure 5, show a significant decrease in the daily (Figure 5 (a)) and cumulative (Figure 5 (b)) COVID-19 cases with increasing vaccination coverage of fully-vaccinated individuals (in relation to the baseline value of the vaccination coverage), as expected. For instance, this figure shows that increasing the baseline value of the fully-vaccinated rate (*ξ*_*v f*_) by 20% resulted in a 12% reduction in daily cases at the peak (Figure 5 (a), gold curve), in comparison to the baseline scenario (Figure 5 (a), blue curve). Further reduction (at least 23% at the peak) is achieved if the baseline value of the fully-vaccinated vaccination coverage is increased by 40% (Figure 5 (a), green curve). On the other hand, if the baseline value of the fully-vaccinated vaccination coverage is decreased, for instance, by 20%, the daily cases at the peak increases (compare magenta curve with blue curve of Figure 5 (a)). Similar reductions in cumulative number of cases are recorded with increasing fully-vaccinated vaccination coverage rate (Figure 5 (b)). In summary, Figure 5 shows that both the daily and cumulative COVID-19 cases can be significantly decreased with even a relatively small increase in the baseline value of the fully-vaccinated vaccination rate (e.g., a 20% increase from baseline coverage rate of the fully-vaccinated coverage rate). This figure also showed (as of the time of writing in March 2022) that the COVID-19 pandemic can be eliminated in the United States, under the baseline vaccination coverage scenario, in late July of 2024. The time-to-elimination is accelerated with increasing values of the baseline fully-vaccinated coverage rate. For instance, Figure 5 (a) showed (as of the time of writing in March 2022) that elimination can be achieved in June of 2022 if the baseline fully-vaccinated coverage rate was increased by 20%.

**Fig. 5:**
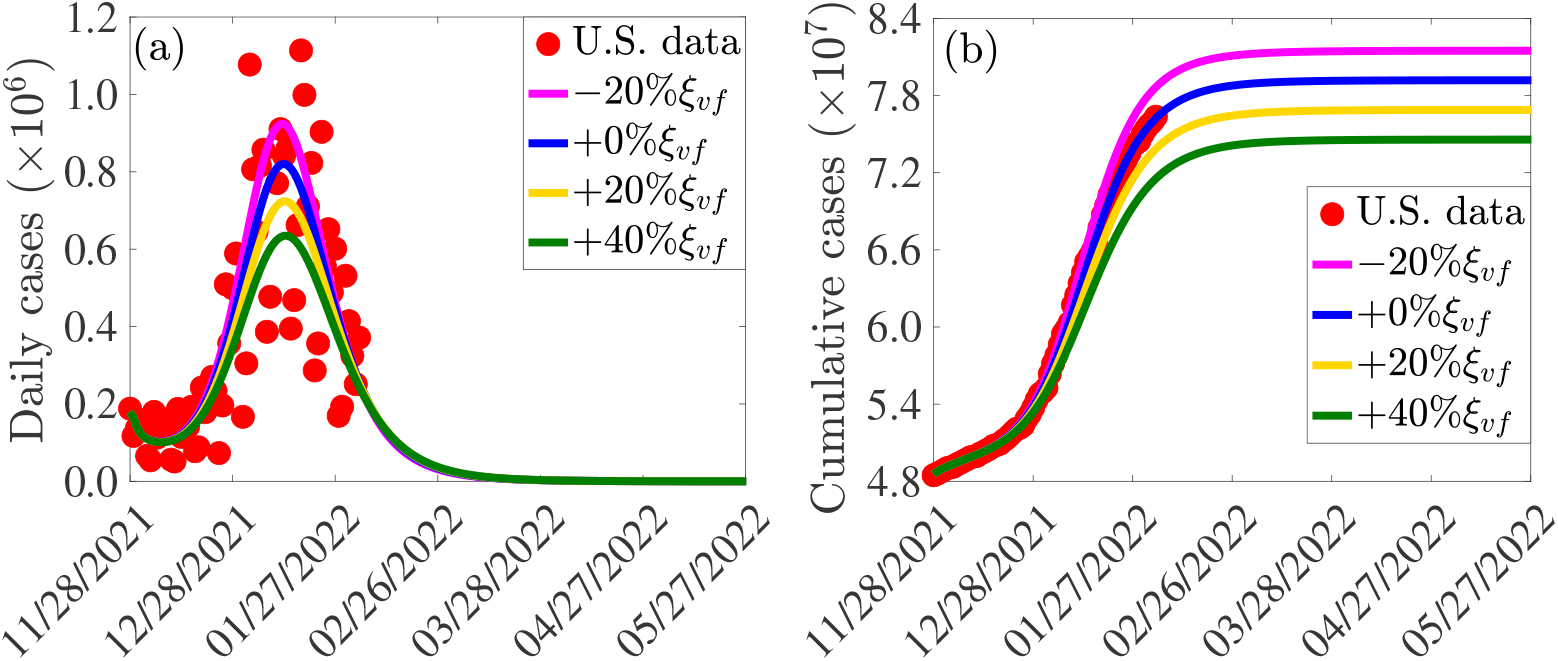
Simulations of the model (2.8) showing the effect of increases or decreases in fully-vaccinated vaccination coverage rate (*ξ*_*v f*_) on the COVID-19 pandemic in the United States. (a) Daily cases, as a function of time, for various values of the fully-vaccinated vaccination coverage rate. (b) Cumulative cases, as a function of time, for various values of the fully-vaccinated vaccination coverage rate. The values of all other parameters used in these simulations are given by the baseline values in Tables S3 and S4.

#### 4.1.1. Assessing the Impact of Additional Increase in Mask Usage from Baseline

The incremental impact of masking coverage (and type) on the effectiveness of the vaccination program (here, too, no treatment of symptomatic individuals is allowed). The simulations are carried out using baseline values of the parameters in Tables S3 and S4, with various values of mask coverage (*c*_*m*_) and types: cloth masks, with estimated efficacy of 30% (i.e., *ε*_*m*_ = 0.3); surgical masks, with estimated efficacy of 70% (i.e., *ε*_*m*_ = 0.7); and N95 masks, with estimated efficacy of 95% (i.e., *ε*_*m*_ = 0.95). The results obtained, depicted in Figure 6, show that, for the fully-vaccinated vaccination coverage (using Pfizer/Moderna vaccine) maintained at the baseline value, increasing coverage of mask usage in the community (*c*_*m*_) resulted in a dramatic reduction in daily COVID-19 cases, particularly if the moderately-effective surgical or the highly-effective N95 masks are prioritized (Figure 6 (a)). For example, under this scenario (with fully-vaccinated vaccination coverage kept at baseline), a 10% increase in mask coverage with surgical mask (Figure 6 (a), dashed gold curve) or N95 respirator (Figure 6 (a), dashed green curve) will result in a 26% and 35% decrease in peak daily cases,respectively, in comparison to the baseline (Figure 6 (a), blue curve). Further reductions are recorded if the coverages of the two mask types are increased by 20% (Figure 6 (a), solid gold and solid green curves). For the case where only the ineffective cloth masks are priorities, a 10% increase in baseline coverage of these masks (Figure 6 (a), dashed magenta curve) will result in approximately 11% decrease in daily cases at the peak, in comparison to the baseline. A further increase to 20% coverage from baseline will result in a 22% decrease in the peak daily cases at baseline (Figure 6 (a), solid magenta curve). Finally, similar reductions in cumulative cases are also recorded with increasing values of the baseline coverage of each of the mask type used in the community (Figure 6 (b)).

**Fig. 6:**
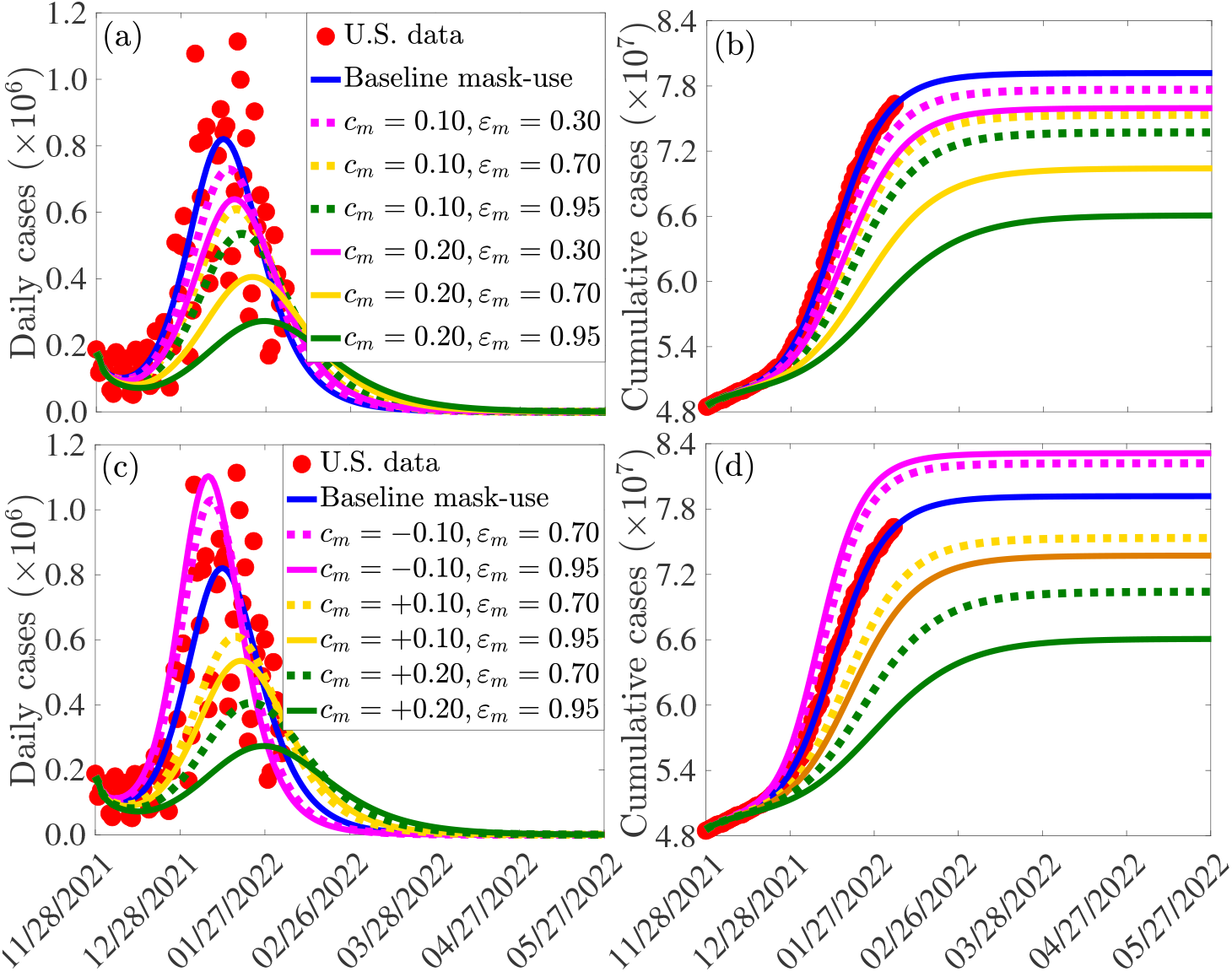
Simulations of the model (2.8) showing the incremental impact of mask coverage (*c*_*m*_) and mask type (cloth masks, with *ε*_*m*_ = 0.3; surgical masks, with *ε*_*m*_ = 0.7; and N95 respirators, with *ε*_*m*_ = 0.95) on the daily ((a) and (c)) and cumulative ((b) and (d)) COVID-19 cases in the United States, as a function of time. The values of the other parameters used in these simulations are as given in Tables S3 and S4.

It should be noted that, in all of the simulations carried out in this subsection (and with the same increase in mask coverage, *c*_*m*_), greater reductions are recorded using N95 masks, followed by surgical mask, and then cloth masks (as expected). Furthermore, our results show that a 20% increase in the baseline value of surgical mask coverage (Figures 6 (a)-(b), solid gold curve) is more effective (in reducing cases) than a 10% increase in baseline N95 coverage (Figures 6 (a)-(b), dashed green curve). In other words, our simulations show that, having more people wear the moderately-effective surgical mask is more effective than having fewer people wear the highly-effective N95 masks. However, this result, which is consistent with that reported in [77] does not hold when N95 or surgical mask is compared with the low-effective cloth mask. Specifically, a 20% increase in the baseline coverage of cloth mask (Figures 6 (a)-(b), solid magenta curve) is not more effective than a 10% increase in the baseline coverage of either surgical (Figures 6 (a)-(b), dashed gold curve) or N95 (Figures 6 (a)-(b), dashed green curve) mask.

Finally, it should be mentioned that relaxing mask usage from the current baseline (as is currently the case in some jurisdictions in the United States partially or fully relaxing mask mandates [78–82]) will result in a re-bounce in disease burden. For example, for the case where mask coverage is decreased by 10% from the current baseline level, our simulations show that a 4-5% increase in the peak daily cases will be recorded if either the surgical mask (Figure 6(c), dashed purple curve) or N95 mask (6(c), solid magenta curve) is prioritized. Similar increases in the cumulative COVID-19 cases are recorded with a decrease in baseline coverage of each of the mask type used in the community (Figure 6 (d)). Thus, our simulations show that, based on the current data and baseline levels of COVID-19 interventions implemented in the United States, relaxing mask mandates will result in increase in new cases (up to about 5% if surgical or N95 are prioritized). Furthermore, giving up masking with surgical mask by a certain proportion is less detrimental than giving up masking with N95 mask by the same proportion (in other words, reducing coverage of surgical mask by 10%, for instance, is less detrimental to the community than reducing the coverage of N95 mask by 10%). This figure also shows that fewer people giving up N95 masks (e.g., 5%) is less detrimental than more people (e.g., 10%) giving up surgical masks.

In summary, the simulations in this subsection show that, for the case where fully-vaccinated vaccination coverage is maintained at baseline level (and no treatment strategy is implemented), increasing the baseline value of mask coverage reduces the daily and cumulative COVID-19 cases, and the level of reduction increases with increasing quality of the mask type that is prioritized in the community. For communities that prioritize the use of the moderately-effective surgical and the highly-effective N95 masks only (i.e., communities that discourage the use of cloth masks, which are known to be generally ineffective [83–86]), having more people (e.g., 20% increase from baseline level) wear surgical masks is more beneficial to the community than having fewer people (e.g., 10% increase from baseline) wear N95 masks. In other words, in this context (with only surgical and N95 masks available), significant increase in the coverage of surgical masks (from their baseline level) may be more effective than a small increase in the baseline coverage of N95 masks.

#### 4.1.2. Assessing the Combined Impact of Vaccination and Mask Usage

The model (2.8) is further simulated to assess the combined impacts of mask coverage (*c*_*m*_), mask type (cloth masks, with *ε*_*m*_ = 0.3; surgical masks, with *ε*_*m*_ = 0.7; and N95 respirators, with *ε*_*m*_ = 0,95) and fully-vaccinated vaccination coverage rate (*ξ*_*v f*_) on the daily and cumulative number of COVID-19 cases in the United States. For these simulations, we consider a 20% increase in the baseline values of both mask coverage (*c*_*m*_) and fully-vaccinated vaccination coverage rate (*ξ*_*v f*_). The results obtained, depicted in Figure 7, show that using the ineffective cloth masks will result to a 33% reduction in the baseline peak daily cases (Figure 7 (a); magenta vs. blue curves). This reduction is better than the 22% reduction in peak daily cases for the scenario where only the baseline vaccination coverage was increased (Figure 6(a), solid magenta curve). The reduction in the number of cases is more significant if masks of higher quality are prioritized. Specifically, if the moderately-effective surgical masks are prioritized, the simulation show that up to 60% reduction in peak daily cases can be achieved (Figure 7 (a), gold curve). The reduction increases to 74% if the highly-effective N95 masks are prioritized (Figure 7 (a), green curve). These reductions exceed the 51% (67%) reductions recorded, for the corresponding scenarios, when the fully-vaccinated vaccination coverage was maintained at its baseline value (Figure 6 (a), solid gold and green curves). Similar reductions in the cumulative cases are recorded with increasing baseline vaccination and mask coverage for each of the mask types used in the community (Figure 7 (b)).

**Fig. 7:**
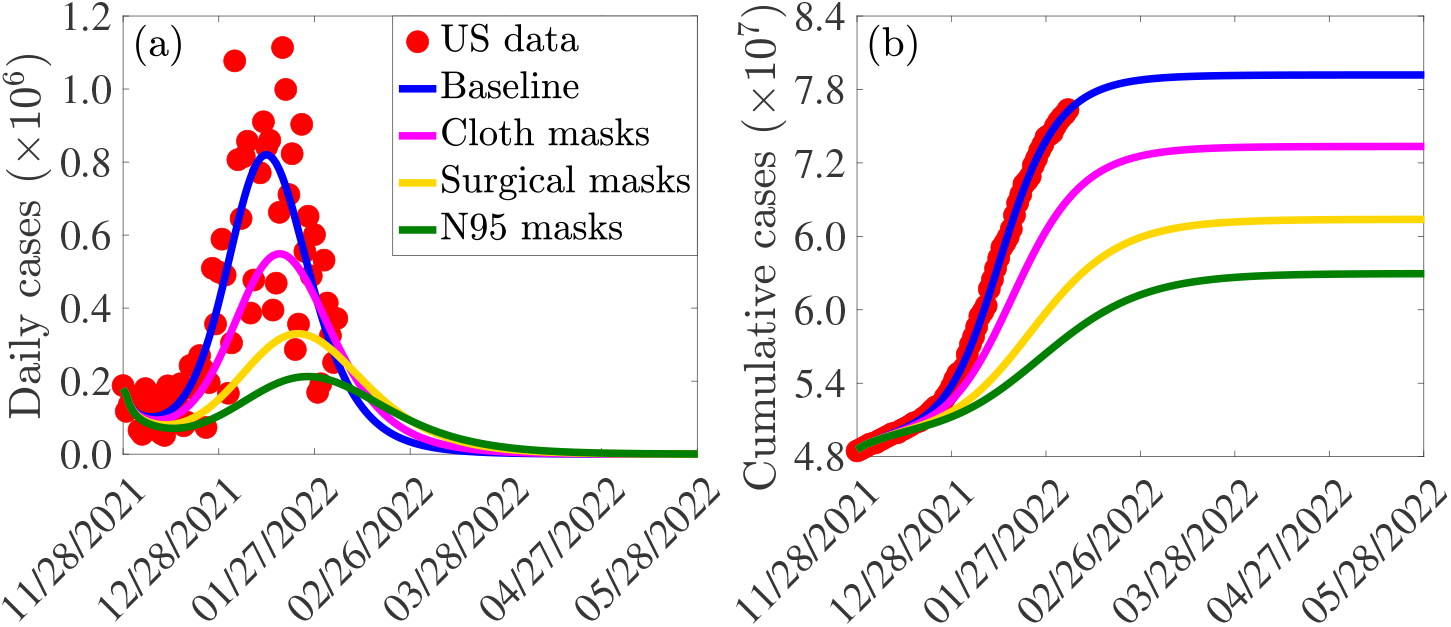
Simulations of the model (2.8), showing the combined incremental impact of mask coverage (*c*_*m*_), mask type (cloth masks, with *ε*_*m*_ = 0.3; surgical masks, with *ε*_*m*_ = 0.7; and N95 respirators, with *ε*_*m*_ = 0,95) and fully-vaccinated vaccination coverage rate (*ξ*_*v f*_) on the daily ((a)) and cumulative ((b)) COVID-19 cases in the United States, as a function of time. In these simulations, the mask and fully-vaccinated vaccination coverage rates are increased by 20% from their respective baseline values. The values of the other parameters used in the simulations are given in Tables S3 and S4.

Contour plots of the control reproduction number (ℝ_*c*_) of the model (2.8), as a function of the vaccine rate coverage (*ξ*_*v f*_) and additional mask coverage (*c*_*m*_) for different mask types, are generated to determine the optimal combinations of the mask and vaccination coverage that can reduce the reproduction number to a value less than unity (so that the pandemic can be eliminated). The results obtained, depicted in Figure S1 in the SI, show that, for a community that prioritizes surgical masks (and the coverage in its usage is held at baseline), the reproduction number can be brought down to a value less than unity if up to 798,000 individuals become fully-vaccinated *per* day (i.e., if the fully-vaccinated vaccination rate is *ξ*_*v f*_ = 0.00532 *per* day; it should be clarified that the number of fully-vaccinated individuals each day is given by *ξ*_*v f*_ *S*.) If the baseline coverage of the surgical mask is increased by 20%, the reproduction number can be brought to a value less than one if 546,000 individuals are fully-vaccinated each day (i.e., if *ξ*_*v f*_ = 0.00364). However, if surgical mask coverage can be increased by 50% from its baseline level, only about 220,500 individuals need to be fully-vaccinated *per* day to bring the reproduction number to a value less than one. On the other hand, if N95 masks are prioritized, the requirement for high daily vaccination rate decreases. For instance, if the N95 coverage is increased by 20% from its baseline, only about 462,000 individuals need to be fully-vaccinated every day to bring the reproduction number to a value less than one (i.e., we need *ξ*_*v f*_ = 0.00308 *per* day, under this scenario). If the baseline coverage of N95 in the community can be increased by 50%, the number of individuals that need to be fully-vaccinated everyday to reduce the reproduction number below one dramatically reduces to 49,500 (i.e., *ξ*_*v f*_ = 0.00033 *per* day in this case). In summary, the contour plots in Figure S1 show that the prospects of eliminating COVID-19 in the United States is more promising if increases in baseline levels of fully-vaccinated vaccination coverage are combined with increases in baseline levels of face mask coverage that prioritizes moderate (surgical) or high quality (N95) masks.

### 4.2. Assessing the Impact of Waning of Vaccine-derived and Natural Immunity

In this section, the model (2.8) is simulated to assess the effect of waning vaccine-derived and natural immunity on the dynamics of the SARS-CoV-2 pandemic in the United States. We first considered the case where only the waning rate of vaccine-derived immunity (for both fully-vaccinated and boosted individuals) varies from its baseline value, while the waning rate of natural immunity remains at its baseline value (of 9 months). If vaccine-derived immunity wanes within 3 months (i.e., *ω*_*v*_ = *ω*_*v f*_ = *ω*_*vb*_ = 1/(0.25 × 365) = 0.011 *per* day), our simulations show that the peak daily cases increases by 8%, in comparison to the case where vaccine-derived immunity wanes at the baseline value of 9 months (Figure 8 (a), magenta curve, in comparison to blue curve). A lower peak of the daily cases is recorded if vaccine-derived immunity wanes in 6 months (Figure 8 (a), gold curve, in comparison to blue curve). If, on the other hand, it takes 4 years for vaccine-derived immunity to wane, our simulations show a marginal decrease in the peak daily cases (about 4% decrease), in comparison to the baseline (Figure 8 (a), green curve, in comparison to blue curve). Similar trends are observed if only waning natural immunity is varied, while the waning rates of vaccine-derived immunity (for both fully-vaccinated and boosted individuals) are maintained at their baseline values (Figure 8 (b)) and when both vaccine-derived and natural immunity are varied from their respective baseline values (Figure 8 (c)). Further, same trends are observed with respect to the cumulative cases for each of the three variable waning rate scenarios (Figures 8 (d)-(f)).

**Fig. 8:**
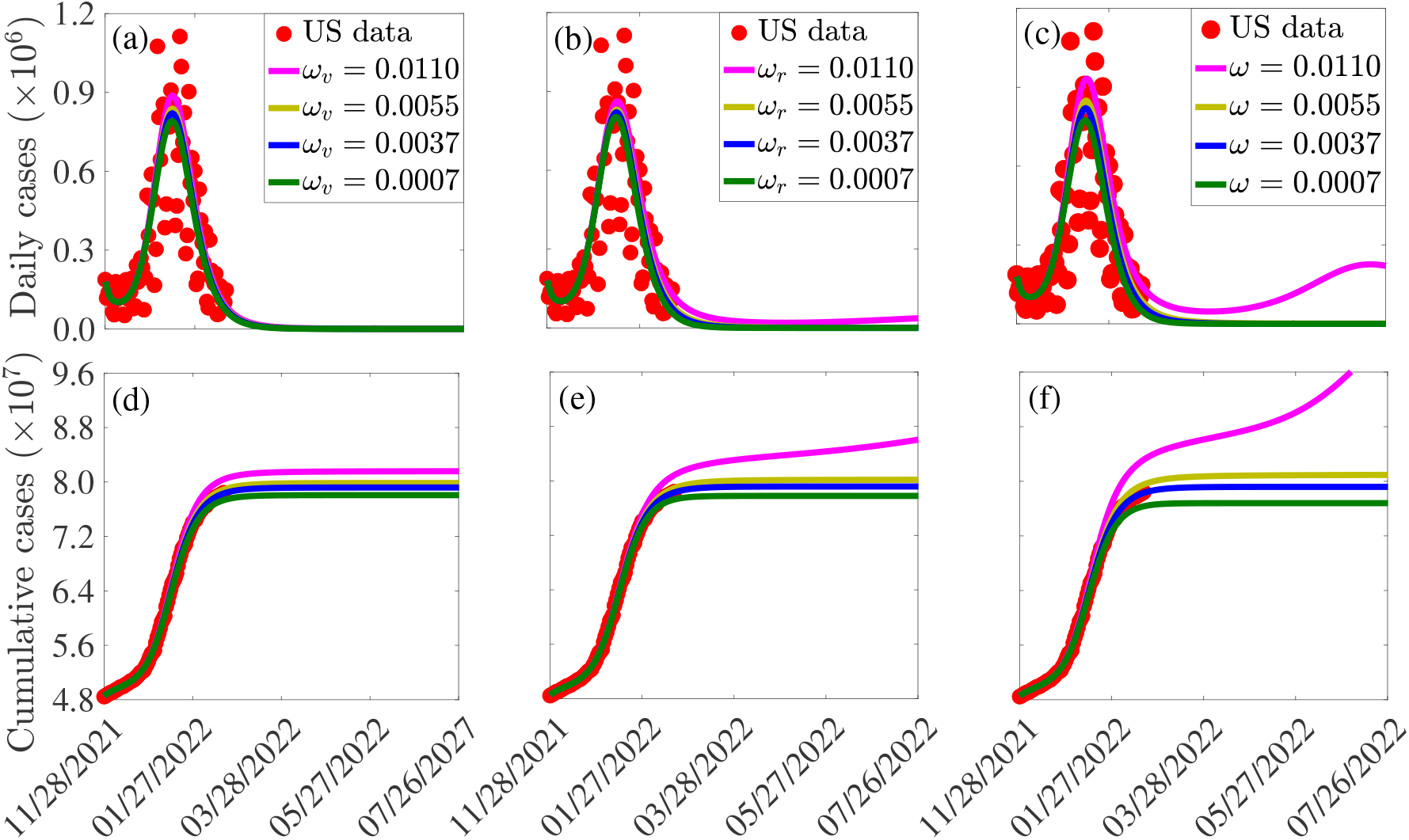
Simulations of the model (2.8), showing (a)-(c) new daily and (d)-(f) cumulative COVID-19 cases in the United States, as a function of time, for various values of the waning rate of (a) and (d) vaccine-derived immunity in fully-vaccinated and boosted individuals (*ω*_*v*_ = *ω*_*v f*_ = *ω*_*vb*_), (b) and (e) natural immunity for the Delta and Omicron variants (*ω*_*r*_ = *ω*_*dr*_ = *ω*_*or*_), and (c) and (f) both vaccine-derived and natural immunity (*ω* = *ω*_*vr*_ = *ω*_*v f*_ = *ω*_*vb*_ = *ω*_*dr*_ = *ω*_*or*_). The durations for the waning of immunity were taken to be 3 months (i.e., *ω* = 0.0110) *per* day), 6 months (*ω* = 0.0055 *per* day), 9 months (*ω* = 0.0037 *per* day) and 48 months (*ω* = 0.0007 *per* day), respectively. The values of the other parameters used in these simulations are as given in Tables S3-S4.

It is worth mentioning that it can be seen from the simulations depicted in Figure 8 that the increase or decrease in disease burden (i.e., daily or cumulative cases) recorded when the waning rates of both the vaccine-derived and natural immunity are varied exceed that for the scenario where only the vaccine-derived or the natural immunity is allowed to vary from baseline. In particular, for the case where both the vaccine-derived and natural immunity wane within 3 months (i.e., *ω* = *ω*_*v f*_ = *ω*_*vb*_ = *ω*_*dr*_ = *ω*_*or*_ = 0.0110 *per* day), a 14% increase in the peak new daily cases is recorded, in comparison to the baseline (we record 8% increase when only vaccine-derived immunity is allowed to vary, as mentioned above). Furthermore, for this case, our simulations suggest (as of the time of writing in March 2022) that another wave of the pandemic that peaks by mid July 2022 (with peak new daily cases about 72% lower than those for the baseline scenario) is predicted to occur (Figure 8 (c), magenta curve). It should, however, be stressed that the simulations in Figure 8 suggest that decreasing waning rate of the vaccine-derived immunity (up to about 4 years) seems to have only a marginal impact in decreasing COVID-19 cases. This may be due to the fact that the two vaccines (Pfizer/Moderna) are highly efficacious against the original SARS-CoV-2 strain, and also moderately-efficacious in the level of cross-protection they offer against the Delta and Omicron variants.

### 4.3. Assessing the Impact of Antiviral Treatment Against COVID-19

In this section, the model (2.8) is simulated to assess the potential impact of the two antiviral drugs (*Paxlovid* and *Molnupiravir*) that received FDA Emergency Use Authorization for use in the United States to treat individuals with clinical symptoms of COVID-19. As of the time of writing (February 2022), the two antivirals are not widely deployed in the United States. Consequently, there isn’t real data to realistically estimate the treatment rates. It should be recalled that in the model (2.8), treatment is offered to symptomatic individuals (during the first five days or after the first five days of onset of symptoms) and hospitalized individuals at rates *τ*_*jk*_ (where *j* = {*d, o*}, representing the two variants; and *k* = 1,2, representing the two symptomatic compartments *I* _*j*1_ and *I* _*j*2_). In the absence of the aforementioned data, our simulations will be carried out for the special case where the treatment rate is the same for each treated compartment (i.e., *τ*_*j*1_ = *τ*_*j*2_ = *τ*_*jh*_ = *τ*, with *j* = {*d,o*}). The simulation results obtained are depicted in Figure 9. This figure shows, first of all, that treatment seems to only have marginal impact in reducing daily new cases (Figure 9 (a)). For instance, if *τ* = 0.8 (i.e., if symptomatic individuals are offered treatment with any of the two antivirals within 1/0.8 = 1.25 days on average), the reduction in daily cases, in comparison to the baseline case is marginal (compare blue and green curves in Figure 9(a)).

**Fig. 9:**
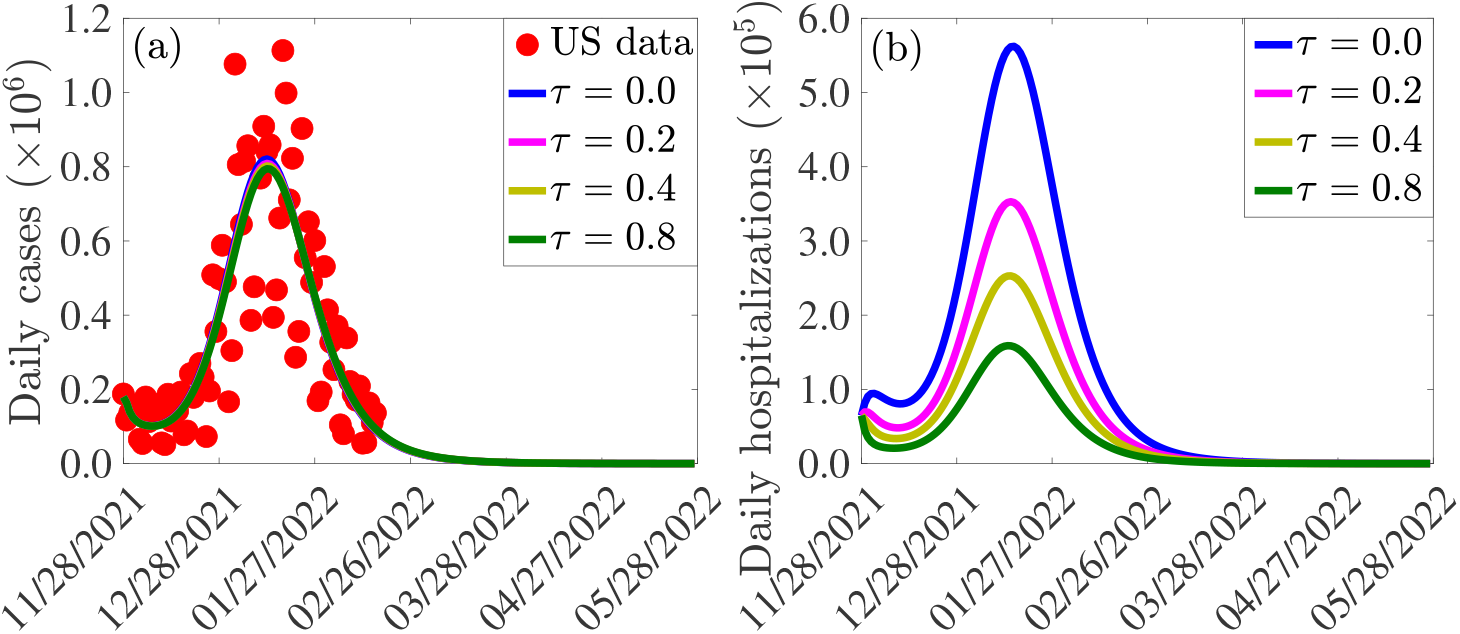
Simulations of the model (2.8) depicting the impact of treatment of symptomatic infectious and hospitalized individuals infectious on the (a) confirmed daily COVID-19 cases, and (b) daily COVID-19 hospitalizations in the United States. The treatment rate (*τ*) is given by *τ* = *τ*_*j*1_ = *τ*_*j*2_ = *τ*_*jh*_, *j* ∈ {*d,o*}. The other parameter values used for the simulations are presented in Tables S3 and S4.

On the other hand, our simulations show that treatment has a more significant impact on reducing daily hospitalization (Figure 9 (b)). For instance, if it takes an average five days to treat a symptomatic individual (i.e., *τ* = 0.2), a 37% reduction in the peak hospitalization will be recorded, in comparison to the baseline (Figure 9(b), compare magenta and blue curves). Under this scenario, and with all other intervention-related parameters kept at their baseline values (as of the time of writing in March 2022), it will take until June 30, 2024 before the number of COVID-related hospitalizations can be significantly reduced to a few or no hospitalizations at all.

If the treatment rate is increased to *τ* = 0.4 (i.e., if it takes an average of 2.5 days to treat a symptomatic individual), the reduction in baseline peak hospitalization increases to 55% (Figure 9(b), compare gold and blue curves). In this case (and as of the time of writing in March 2022), zero hospitalization (i.e., as stated above, reducing the number of individuals that are hospitalized with the disease to essentially a few or no cases), can be achieved by June 19, 2024. Finally, the reduction in baseline peak hospitalization increases to 72% if *τ* = 0.8 (Figure 9(b), compare green and blue curves). For this scenario, zero hospitalization can be achieved by June 6, 2024 (this will be visible if Figure 9 is extended to 2024). In summary, the simulations in this subsection show that while treatment only has marginal impact on reducing number of cases, it does have significant impact in reducing daily hospitalizations. With all other interventions (vaccination and masking) maintained at their baseline values, our simulations show that treatment, even at perhaps the highest possible rate (such as a rate associated with treating symptomatic individuals within a day or two of onset of symptoms) will delay COVID-19 elimination (as measured, in these simulations, in terms of attaining zero hospitalization) until 2024. Such elimination can be achieved by increasing the effectiveness and coverages of vaccination and/or mask usage. In other words, the simulations suggest that, while treatment reduces hospitalization (a highly desirable goal too), the prospect of COVID-19 elimination is enhanced by focusing investments on mask usage and vaccination than on treatment.

## 5. Discussion and Conclusions

One of the main success stories in the effort to control the spread of the devastating novel 2019 coronavirus pandemic (COVID-19, caused by SARS-CoV-2) was the rapid development and deployment of safe and effective vaccines [87–90]. In particular, the United States Food and Drug Administration (FDA) approved three such vaccines for use in the United States, namely the Pfizer-BioNTech, Moderna and Johnson & Johnson vaccines. Prior to the game-changing moment of the development of these vaccines, control efforts against the SARS-CoV-2 pandemic were limited to the use of non-pharmaceutical interventions, such as quarantine of suspected case, isolation of confirmed cases, use of face masks in public, social-distancing, community lockdowns, etc. Although the vaccines have proven to be very effective in significantly decreasing the burden of the SARS-CoV-2 pandemic in the United States and in other countries and/or regions with moderate to high vaccination coverage (as measured in terms of preventing new cases and minimizing severity of disease, hospitalization and death in breakthrough infections), they have not been able to lead to the elimination of the pandemic. This is largely due to the concerning level of vaccine hesitancy [21, 91–98] and emergence of numerous variants of concern [35–40, 42, 43]. The three vaccines being used in the United States were designed to target the original strain of SARS-CoV-2, and they only offer cross-protective efficacy against the variants that emerged and circulate in the population. The FDA also provided Emergency Use Authorization (EUA) for two very effective antivirals for use to treat individuals with severe symptoms of SARS-CoV-2.

Since it is now clear that vaccination alone is insufficient to lead to elimination of the pandemic, it is instructive to develop and use a mathematical modeling framework for assessing the community-wide impact of combining the vaccination program with other control strategies (NPIs, such as face mask usage, and the two antivirals) in an effort to effectively curtail the COVID-19 pandemic in the United States, and for the scenario where two variants of concern (Delta and Omicron) are co-circulating. This forms the objective of this study. We developed a mathematical model, of the form of a deterministic system of nonlinear differential equations, for assessing the combined impacts of vaccination, face mask usage and antiviral treatment on minimizing and mitigating the burden of the two COVID-19 variants. In our model formulation, vaccination is based on using either the Pfizer or Moderna vaccine (each with protective efficacy against acquisition of infection set at about 95%). We fitted and parameterized the model using the observed daily COVID-19 case data for the United States starting from the time when Omicron first emerged (end of November 2021) to the end of January 2022. We then used the additional available data for the period from February 1, 2022 to March 10, 2022 for cross validation purpose (i.e., to validate the model). The cross-validation showed that the model perfectly predicts the case data for the time period from February 1, 2022 to March 8, 2022.

Qualitative analysis of the model reveals that its disease-free equilibrium is locally-asymptotically stable whenever a certain epidemiological threshold, known as the *control reproduction number* (denoted by ℝ_*c*_) is less than one. This result is extended to establish the global asymptotic stability of the disease-free equilibrium for a special case of the model. The epidemiological implication of this asymptotic stability result for the disease-free equilibrium of the model is that the SARS-CoV-2 pandemic can be eliminated in the United States if the control measures implemented can bring (and maintain) the reproduction number to a value less than one. The pandemic will persist if the control measures are unable to bring the control reproduction number to a value less than one.

We computed, using the baseline values of the parameters of our model (which was parameterized using observed daily case data for the pandemic in the United States), the value of the threshold quantity ℝ_*c*_, which is expressed as the maximum of the constituent reproduction numbers for the spread of the Delta and Omicron variants. This computation showed that, for the simulation period from November 28, 2021 to March 1, 2022, the constituent reproduction number for the Delta variant is 0.28 while that for the Omicron variant is approximately equal to one (0.96). Thus, our study show that Omicron is the predominant variant, and that Delta has essentially died out. Furthermore, if current baseline levels of the control measures implemented in the United States are maintained, Omicron will die out as well (but will persist if the control measures are relaxed, to the extent that the reproduction number for Omicron exceeds one).

This study further showed that vaccine-derived herd immunity can be achieved (and the pandemic can be eliminated) if at least 68% of the population is fully-vaccinated with either the Pfizer or Moderna vaccine. Since data from the CDC shows, as of February 14, 2022 that 64% of United States population is already fully-vaccinated [75, 76], our study suggests that increasing the vaccination coverage in the unvaccinated population (and/or population of those who only received one vaccine dose) by about 4% could push the population to achieve herd immunity. If the level of cross-protective immunity offered by the two vaccines against the Delta and Omicron variants is lower than the baseline levels in our simulations (e.g., 60%), the vaccination coverage needed to achieve vaccine-derived herd immunity increases (to 84%).

We also assessed the impact of combining vaccination with mask usage using various mask types. We showed that the proportion of individuals who need to be fully-vaccinated to achieve herd immunity decreases with increasing coverage of face masks in the community (from the baseline face mask usage), and the level of reduction achieved depends on the quality of the mask used (specifically, a greater reduction in herd immunity level needed to eliminate the disease is achieved if the high-quality N95 masks are prioritized, in comparison to the scenario where moderate quality surgical masks are prioritized). We showed that greater reductions in pandemic burden are recorded using N95 masks, followed by surgical masks, and then cloth masks (as expected). Furthermore, our results show that having more people wear surgical masks is more effective than fewer people wearing N95 masks. This result indicates that the SARS-CoV-2 pandemic would have been easily controlled if more surgical masks (which are moderately-effective) were made available to the populace during the early stages of the pandemic (since this would have slowed down the spread of the SARS-CoV-2 pandemic, while the three safe and effective vaccines authorized for use in the United States were developed).

Although the mRNA vaccines (Pfizer, Moderna) designed to fight the original strain of SARS-CoV-2 reduce the risk of hospitalization and death and offer some cross protection against variants of concern, the cross protective efficacy they offer wane over time. In particular, the protective efficacy of these vaccines wanes down to about 40% within a few months of the second dose [17, 99, 100]. We simulated the model to assess the impact of waning vaccine-derived immunity (in fully vaccinated and boosted individuals) and natural immunity. Our simulations showed an increase in disease burden if immunity wanes at a faster rate (e.g., if both vaccine-derived and natural immunity wane within 3 months, as against the baseline of within about 9 months to a year), with the possibility of another wave of the Omicron variant (*albeit* a much milder one this time, with a projected peak size at least 72% lower than the Omicron peak size recorded in January of 2022). This, and the possibility of future emergence of other SARS-CoV-2 variants of concern (to compete with, or displace, Omicron), suggests that a fourth Pfizer/Moderna booster dose may be needed in the United States this year (2022) to supplement the effort to eliminate the SARS-CoV-2 pandemic. For example, laboratory investigations have suggested that the BA.2 sub-variant of Omicron might lead to more severe disease, and that current vaccines against COVID-19 might not be effective against this sub-variant [101–103]. In any case, our simulation results advocating for a fourth booster dose of Pfizer and Moderna for the United States, is in line with the decision in Israel to authorize a fourth booster dose against the pandemic starting with immuno-compromised individuals, adults over the age of 60 and health-care employees, and then adults aged 18 and over [104, 105]. It should be emphasized that our simulations show only a marginal increase in disease burden if both vaccine-derived and natural immunity last at least 6 months.

We showed that while the use of the two approved antiviral drugs induce marginal impact in reducing the number of new daily cases of SARS-CoV-2 in the United States, their usage offer a more pronounced effect in reducing hospitalizations. Specifically, if all other interventions (vaccination and masking) are maintained at their baseline values, even the most efficient treatment strategy (e.g., one associated with treating symptomatic cases within a day or two of onset of symptoms) cannot lead to elimination of the COVID-19 pandemic until about the year 2024. However, such elimination can be achieved by increasing the effectiveness and coverages of vaccination and/or mask usage. In other words, our study showed that while treatment reduces hospitalizations, the prospect of COVID-19 elimination is enhanced by focusing investments of control resources on mask usage and vaccination, rather than on treatment options.

## Data Availability

All data produced in the present study are available upon reasonable request to the authors

## 6. Acknowledgments

ABG acknowledges the support, in part, of the Simons Foundation (Award #585022) and the National Science Foundation (Grant Number: DMS-2052363). CNN acknowledges the support of the Simons Foundation (Award #627346) and the National Science Foundation (Grant Number: DMS #2151870). HBT acknowledges the support of the Graduate Research Assistantship in Developing Countries Program (from the International Mathematics Union) and Centre d’Excellence Africain en Sciences Mathematiques, Informatique et Applications, Benin. SS acknowledges the support of the Fulbright Foreign Student Program.

## 7. Additional information

### 7.2. Data availability

The data sets used and/or analysed during the current study are available from the websites:

- https://github.com/CSSEGISandData/COVID-19
- https://ourworldindata.org/coronavirus/country/united-states

Relevant codes used for analyzing the data and for producing the figures in the paper are uploaded on the appropriate journal platform.

