## Supplementary information for "Unraveling the dynamics of the Omicron and Delta variants of the 2019 coronavirus in the presence of vaccination, mask usage, and antiviral treatment"

#### Online Supplementary Information

#### S1 Description of the State Variables and Parameters of the Model

Table S1: Description of the state variables of the model (2.8). The subscript $j = d$ denotes the Delta variant and $j = o$ denotes the Omicron variant.

| State variables | Description |
|---|---|
| $S$ | Population of unvaccinated susceptible individuals (and individuals who lost their immunity over time) |
| $V_f$ | Population of fully-vaccinated but not boosted individuals |
| $V_b$ | Population of fully-vaccinated individuals who received the booster dose |
| $E_j$ | Population of exposed (i.e., newly-infected or latent) individuals |
| $P_j$ | Population of pre-symptomatically infectious individuals |
| $A_j$ | Population of asymptomatically-infectious individuals |
| $Q_j$ | Population of detected (positive) exposed, presymptomatic and asymptomatic individuals |
| $I_{j1}$ | Population of symptomatically-infectious individuals within the first five days of the onset of SARS-CoV-2 symptoms |
| $I_{j2}$ | Population of symptomatically-infectious individuals that survived the first five days of the onset of SARS-CoV-2 symptoms |
| $H_j$ | Population of hospitalized individuals |
| $R_j$ | Population of recovered and successfully treated individuals |
| $N$ | Total population ($N = S + V_f + V_b + E_d + P_d + A_d + Q_d + I_{d1} + I_{d2} + H_d + R_d + +E_o + P_o + A_o + Q_o + I_{o1} + I_{o2} + H_o + R_o$) |

Table S2: Description of the parameters of the model (2.8). Notation: $j \in \{d, o\}$, where the subscript $d$ denotes the Delta variant and the subscript $o$ denotes the Omicron variant.

| **Parameter** | **Description** |
|---|---|
| $\Lambda$ | Recruitment rate (by birth or immigration) into the population |
| $\mu$ | Natural death rate |
| $\beta_{jk}$ | Effective contact rate for infectious individuals in $P_j$ class ($k = p$), $A_j$ class ($k = a$), $I_{jk}$ class ($k = 1, 2$), and the $H_j$ class ($k = h$) |
| $q_1$ | Modification parameter accounting for the proportion of individuals in the $Q_j$ class who are actually infectious |
| $\xi_{vf}$ | Rate at which susceptible individuals are fully-vaccinated |
| $\xi_{vb}$ | Rate at which fully -Vaccinated individuals receive the booster dose |
| $\varepsilon_{jf}$ | Cross-protective efficacy of the vaccine against acquisition of the Delta ($j = d$) or Omicron ($j = o$) variant for fully-vaccinated but not boosted individuals |
| $\varepsilon_{jb}$ | Cross-protective efficacy of the vaccine against acquisition of the Delta ($j = d$) or Omicron ($j = o$) variant for boosted individuals |
| $\omega_{jf}$ | Rate of waning of vaccine-derived immunity for fully- vaccinated but not boosted individuals |
| $\omega_{jb}$ | Vaccine waning rate for fully-vaccinated and boosted individuals |
| $\omega_{jr}$ | Rate of loss of infection-acquired (natural) immunity |
| $\sigma_{je}$ | Progression rate of exposed humans to the presymptomatically-infectious stage |
| $\sigma_{jp}$ | Progression rate of presymptomatically-infectious individuals at the end of the incubation period to the asymptomatically-infectious ($A_j$) or the first symptomatically-infectious ($I_{d1}$) class |
| $\alpha_{j1}$ | Transition rate of individuals from the $I_{j1}$ to the $I_{j2}$ class (i.e., $1/\alpha_{j1} = 5$ days) |
| $\rho_j$ | Detection or positivity rate of exposed, presymptomatic and asymptomatic individuals |
| $\psi_j$ | Rate at which detected individuals develop symptoms |
| $r_j(1 - r_j)$ | Proportion of presymptomatically-infectious individuals who exhibit (do not exhibit) clinical symptoms of SARS-CoV-2 at the end of the incubation period |
| $\phi_{j1}$ $(\phi_{j2})$ | Hospitalization rate of individuals in the $I_{j1}$ $(I_{j2})$ class |
| $\gamma_{ja}$ $\gamma_{jq}$ $(\gamma_{j2})$ $(\gamma_{jh})$ | Natural recovery rate for individuals in the $A_j$ $(Q_j)$ $(I_{j2})$ $(H_j)$ class |
| $\tau_{j1}$ $(\tau_{j2})$ $(\tau_{jh})$ | Treatment rate of individuals in the $I_{j1}$ $(I_{j2})$ $(H_j)$ class |
| $\delta_{j1}$ $(\delta_{j2})$ $(\delta_{jh})$ | Disease-induced death rate of symptomatically-infectious humans in the $I_{j1}$ $(I_{j2})$ $(H_j)$ class |

#### S2 Baseline Values of Fixed and Estimated (Fitted) Parameters of the Model

Table S3: Baseline values of fixed parameters of the model (2.8). Apart from the vaccine efficacies ($\varepsilon_j$, $\varepsilon_{jk}, j \in \{d, o\}, k \in \{b, f\}$), the proportions $r_j$, and the modification parameter $q_1 = 0.85$, which are dimensionless, all the other parameters have units of *per* day. Notation: $^*$ represents "adapted from" the cited reference.

| Parameter | Value | Source |
|---|---|---|
| $\Lambda$ | 11400 | [1] |
| $\mu$ | $3.4 \times 10^{-5}$ | [1] |
| $\xi_{vf}$ | $1.9 \times 10^{-5}$ | [2] |
| $\xi_{vb}$ | $2.6 \times 10^{-5}$ | [2] |
| $\delta_{dh}$ | $1.0 \times 10^{-4}$ | [1]* |
| $\delta_{oh}$ | $5.0 \times 10^{-5}$ | [1]* |
| $\delta_{d1}$ | $2.5 \times 10^{-5}$ | [1]* |
| $\delta_{o1}$ | $1.3 \times 10^{-5}$ | [1]* |
| $\delta_{d2}$ | $5.0 \times 10^{-5}$ | [1]* |
| $\delta_{02}$ | $2.5 \times 10^{-5}$ | [1]* |
| $\rho_d, \rho_o$ | $1.2 \times 10^{-4}$ | [3] |

| Parameter | Value | Source |
|---|---|---|
| $\alpha_{d1}, \alpha_{o1}$ | $1/5$ | [4] |
| $\omega_{vf}$ | $1/274$ | [5]* |
| $\omega_{vb}$ | $1/365$ | [5]* |
| $\omega_{dr}, \omega_{or}$ | $1/274$ | [5] |
| $r_d, r_o$ | 0.200 | [6, 7] |
| $\varepsilon_{df}$ | 0.950 | [8] |
| $\varepsilon_{of}$ | 0.880 | [9] |
| $\varepsilon_{db}$ | 0.950 | [10] |
| $\varepsilon_{ob}$ | 0.755 | [9] |
| $\sigma_{de}$ | $1/5$ | [11] |
| $\sigma_{oe}$ | $1/3$ | [12] |

| Parameter | Value | Source |
|---|---|---|
| $\sigma_{dp}, \sigma_{op}$ | $1/2$ | [11] |
| $\phi_{d1}, \phi_{o1}$ | $1/5$ | [11]* |
| $\phi_{d2}, \phi_{o2}$ | $1/3$ | [11]* |
| $\gamma_{da}$ | $1/5$ | [13] |
| $\gamma_{oa}$ | $2/5$ | [13]* |
| $\gamma_{dh}, \gamma_{dq}$ | $1/5$ | [13]* |
| $\gamma_{oh}, \gamma_{oq}$ | $2/5$ | [13]* |
| $\gamma_{d2}$ | $1/4$ | [13]* |
| $\gamma_{o2}$ | $2/4$ | [13]* |
| $\psi_d$ | $1/3$ | [11]* |
| $\psi_o$ | $1/3$ | [11, 12]* |

Table S4: Baseline values of the estimated (fitted) parameters (and Confidence Intervals (CIs)) of the model (2.8) using COVID-19 confirmed case data for the United States for the period from November 28, 2021 to February 2, 2022. The unit of each of the estimated community transmission rate is *per* day.

(a) Delta variant related parameters. For this baseline set of parameter values, $\mathbb{R}_{dv} = 0.2782$, with 95% CI $[0.1991, 0.5197]$. The basic reproduction number is $\mathbb{R}_{0d} = 0.6929$, with 95% CI $[0.2914, 1.3668]$.

| Parameter | value | 95% CI |
|---|---|---|
| $\beta_{dp}$ | 0.331690 | [0.14489240, 0.44988790] |
| $\beta_{da}$ | 0.003185 | [0.00024100, 0.13617420] |
| $\beta_{d1}$ | 0.024774 | [0.00151000, 0.18891280] |
| $\beta_{d2}$ | 0.000579 | [0.00000010, 0.00097120] |
| $\beta_{dq}$ | 0.000008 | [0.00000001, 0.00018700] |
| $\beta_{dh}$ | 0.000004 | [0.00000001, 0.00008820] |
| $\xi_{vf}$ | 0.005863 | [0.00179810, 0.00643250] |
| $\xi_{vb}$ | 0.000992 | [0.00030424, 0.00108838] |

(b) Omicron variant related parameters. For this set of parameter values, $\mathbb{R}_{ov} = 0.9602$ with 95% CI $[0.6206, 1.7509]$. The basic reproduction number is $\mathbb{R}_{0o} = 2.0587$ with 95% CI $[0.8732, 3.9008]$.

| Parameter | value | 95% CI |
|---|---|---|
| $\beta_{op}$ | 0.995070 | [0.43467720, 1.34966370] |
| $\beta_{oa}$ | 0.009555 | [0.00072300, 0.40852260] |
| $\beta_{o1}$ | 0.074322 | [0.00453000, 0.56673840] |
| $\beta_{o2}$ | 0.001737 | [0.00000030, 0.00291360] |
| $\beta_{oq}$ | 0.000024 | [0.00000003, 0.00056100] |
| $\beta_{oh}$ | 0.000012 | [0.00000003, 0.00026460] |
| $\xi_{vf}$ | 0.005863 | [0.00179810, 0.00643250] |
| $\xi_{vb}$ | 0.000992 | [0.00030424, 0.00108838] |

#### S3 Proof of Theorem 3.2

In this section, the global asymptotic stability of the disease-free equilibrium of the model (2.8) will be established for the special case with negligible disease-induced mortality (i.e., $\delta_{j1} = \delta_{j2} = \delta_{jh} = 0$; where $j \in \{d, o\}$) and no warning of both the vaccine-derived immunity for fully-vaccinated individuals (i.e., $\omega_{vf} = 0$) and natural immunity (i.e., $\omega_{dr} = \omega_{or} = 0$). Before proving the global asymptotic stability of the model, it is necessary to establish the positive-invariance and attractivity of the region $\Omega_*$, as done below.

##### S3.1 Proof of positive invariance and attractivity of $\Omega_*$

Recall the following feasible region (for the special case of the model)

$$\Omega_* = \left\{(S, V_f, V_b, E_d, P_d, A_d, Q_d, I_{d1}, I_{d2}, H_d, R_d, E_o, P_o, A_o, Q_o, I_{o1}, I_{o2}, H_o, R_o) \in \mathbb{R}^{19}_+ : S \le S^*, V_f \le V_f^*, V_b \le V_b^*\right\},$$

with $S^*$, $V_f^*$, and $V_b^*$ as given in Equation (3.1), but with $\omega_{vf} = 0$. We claim the following result.

**Theorem S3.1.** *The region $\Omega_*$ is positively-invariant and attracts all initial solutions of the special case of the model (2.8).*

*Proof.* It can be seen, by adding all the equations of the special case of the model (2.8), that the equation for the rate of change of the total human population is given by:

$$\frac{dN}{dt} = \Lambda - \mu N,$$

from which it follows that $N(t) \to \Lambda/\mu$ as $t \to \infty$. From now on, we replace $N(t)$ with its limiting value, $N^* = \Lambda/\mu$, in the special case of the model (i.e., the standard incidence formulation for the infection rate is now replaced by a mass action incidence). The proof is based on the approach discussed in [14]. It can be seen from the first equation of the special case of the model (2.8) that:

$$\begin{aligned}
\frac{dS}{dt} &\le \Lambda + \omega_{vb} V_b - (\xi_{vf} + \mu)S, \\
&\le \Lambda + \omega_{vb}\left(\frac{\Lambda}{\mu} - S\right) - (\xi_{vf} + \mu)S \\
&\le \frac{\Lambda(\omega_{vb} + \mu)}{\mu} - (\omega_{vb} + \xi_{vf} + \mu)S, \\
&\le \frac{\Lambda(\omega_{vb} + \mu)(\xi_{vb} + \mu)}{\mu(\xi_{vb} + \mu)} - (\omega_{vb} + \xi_{vf} + \mu)S \\
&\le (\omega_{vb} + \xi_{vf} + \mu)\left[\frac{\Lambda(\omega_{vb} + \mu)(\xi_{vb} + \mu)}{\mu(\omega_{vb} + \xi_{vf} + \mu)(\xi_{vb} + \mu)} - S\right], \\
&\le (\omega_{vb} + \xi_{vf} + \mu)\left[\frac{\Lambda(\omega_{vb} + \mu)(\xi_{vb} + \mu)}{\mu((\xi_{vb} + \mu)(\omega_{vb} + \mu) + \xi_{vf}(\omega_{vb} + \mu) + \xi_{vb}\xi_{vf})} - S\right] \\
&= (\omega_{vb} + \xi_{vf} + \mu)\left(S^* - S\right).
\end{aligned}$$

Hence, if $S(t) > S^*$, then $\frac{dS}{dt}$ is negative. Thus, $S(t) \le S^*$ for all $t$, provided that $S(0) \le S^*$. Using similar approach for the second and the third equations of the special case of the model, and using the above bound, leads to the following bounds:

$$\frac{dV_f}{dt} \le \xi_{vf} S^* - (\xi_{vb} + \mu)V_f = (\xi_{vb} + \mu)(V_f^* - V_f),$$

and,

$$\frac{dV_b}{dt} \le \xi_{vb} V_f^* - (\omega_{vb} + \mu)V_b = (\omega_{vb} + \mu)(V_b^* - V_b).$$

Following the same argument, we have $V_f(t) \le V_f^*$ and $V_b(t) \le V_b^*$ for all $t$, provided that $V_f(0) \le V_f^*$ and $V_b(0) \le V_b^*$. It follows from these bounds that $\Omega_*$ is positively invariant and attracts all initial solutions of the special case of the model (2.8). □

#### S3.2 Next generation matrices for the special case of the model

For the aforementioned special case of the model (2.8), it can be shown that the associated next generation matrices are given, respectively, by (note that, throughout this section, $j \in \{d, o\}$):

$$\hat{F_j} = \begin{bmatrix} 0 & \hat{f_{j1}} & \hat{f_{j2}} & \hat{f_{j3}} & \hat{f_{j4}} & \hat{f_{j5}} & \hat{f_{j6}} \\ 0 & 0 & 0 & 0 & 0 & 0 & 0 \\ 0 & 0 & 0 & 0 & 0 & 0 & 0 \\ 0 & 0 & 0 & 0 & 0 & 0 & 0 \\ 0 & 0 & 0 & 0 & 0 & 0 & 0 \\ 0 & 0 & 0 & 0 & 0 & 0 & 0 \\ 0 & 0 & 0 & 0 & 0 & 0 & 0 \end{bmatrix},$$

and,

$$\hat{V_j} = \begin{bmatrix} \hat{K_{j1}} & 0 & 0 & 0 & 0 & 0 & 0 \\ -\sigma_j e & \hat{K_{j2}} & 0 & 0 & 0 & 0 & 0 \\ 0 & -(1-r_j)\sigma_{jp} & \hat{K_{j3}} & 0 & 0 & 0 & 0 \\ -\rho_j & -\rho_j & -\rho_j & \hat{K_{j4}} & 0 & 0 & 0 \\ 0 & -r_j\sigma_{jp} & 0 & -\psi_j & \hat{K_{j5}} & 0 & 0 \\ 0 & 0 & 0 & 0 & -\alpha_{j1} & \hat{K_{j6}} & 0 \\ 0 & 0 & 0 & 0 & -\phi_{j1} & -\phi_{j2} & \hat{K_{j7}} \end{bmatrix},$$

where (with $S^*$, $V_f^*$ and $V_b^*$ as defined in Equation (3.1), but with $\omega_{vf} = 0$),

$$\begin{aligned} \hat{f_{j1}} &= \beta_{jp}\left(S^* + (1-\varepsilon_{jf})V_f^* + (1-\varepsilon_{jb})V_b^*\right), \hat{f_{j2}} = \beta_{ja}\left(S^* + (1-\varepsilon_{jf})V_f^* + (1-\varepsilon_{jb})V_b^*\right), \\ \hat{f_{j3}} &= \beta_{jq}\left(S^* + (1-\varepsilon_{jf})V_f^* + (1-\varepsilon_{jb})V_b^*\right), \hat{f_{j4}} = \beta_{j1}\left(S^* + (1-\varepsilon_{jf})V_f^* + (1-\varepsilon_{jb})V_b^*\right), \\ \hat{f_{j5}} &= \beta_{j2}\left(S^* + (1-\varepsilon_{jf})V_f^* + (1-\varepsilon_{jb})V_b^*\right), \hat{f_{j6}} = \beta_{jh}\left(S^* + (1-\varepsilon_{jf})V_f^* + (1-\varepsilon_{jb})V_b^*\right), \end{aligned}$$

and,

$$\begin{aligned} \hat{K_{j1}} &= \sigma_{je} + \rho_j + \mu, \hat{K_{j2}} = \sigma_{jp} + \rho_j + \mu, \hat{K_{j3}} = \gamma_{ja} + \rho_j + \mu, \hat{K_{j4}} = \gamma_{j_q} + \psi_j + \mu, \\ \hat{K_{j5}} &= \tau_{j1} + \alpha_{j1} + \phi_{j1} + \mu, \hat{K_{j6}} = \tau_{j2} + \gamma_{j2} + \phi_{j2} + \mu, \text{ and } \hat{K_{j7}} = \tau_{jh} + \gamma_{jh} + \mu. \end{aligned}$$

It is convenient to define the following threshold quantity:

$$\hat{\mathbb{R}}_c = \mathbb{R}_c|_{\delta_{j1}=\delta_{j2}=\delta_{jh}=\omega_{vf}=0} \text{ (with } \mathbb{R}_c \text{ as defined in Equation (3.4)).}$$

#### S3.3 Proof of Theorem 2.3

*Proof.* Consider the special case of the model (2.8) with $\delta_{j1} = \delta_{j2} = \delta_{jh} = 0$ and $\omega_{vf} = \omega_{dr} = \omega_{or} = 0$. Furthermore, let $\hat{\mathbb{R}}_c < 1$. The equations for the infected compartments of this special case of the model can be re-written in terms of the next generation matrices ($\hat{F_j}$ and $\hat{V_j}$, given above) as follows:

$$\frac{d}{dt}\begin{bmatrix} E_j(t) \\ P_j(t) \\ A_j(t) \\ Q_j(t) \\ I_{j1}(t) \\ I_{j2}(t) \\ H_j(t) \end{bmatrix} = (\hat{F_j} - \hat{V_j})\begin{bmatrix} E_j(t) \\ P_j(t) \\ A_j(t) \\ Q_j(t) \\ I_{j1}(t) \\ I_{j2}(t) \\ H_j(t) \end{bmatrix} - \hat{M_j}\begin{bmatrix} E_j(t) \\ P_j(t) \\ A_j(t) \\ Q_j(t) \\ I_{j1}(t) \\ I_{j2}(t) \\ H_j(t) \end{bmatrix}, \tag{S.1}$$

where (with $S^*$, $V_f^*$ and $V_b^*$ as defined in Equation (3.1), but with $\omega_{vf} = 0$),

$$
(\hat{F}_j - \hat{V}_j) = \begin{bmatrix} -\hat{K_{j1}} & \hat{f_{j1}} & \hat{f_{j2}} & \hat{f_{j3}} & \hat{f_{j4}} & \hat{f_{j5}} & \hat{f_{j6}} \\ \sigma_j e & -\hat{K_{j2}} & 0 & 0 & 0 & 0 & 0 \\ 0 & (1-r_j)\sigma_{jp} & -\hat{K_{j3}} & 0 & 0 & 0 & 0 \\ \rho_j & \rho_j & \rho_j & -\hat{K_{j4}} & 0 & 0 & 0 \\ 0 & r_j\sigma_{jp} & 0 & \psi_j & -\hat{K_{j5}} & 0 & 0 \\ 0 & 0 & 0 & 0 & \alpha_{j1} & -\hat{K_{j6}} & 0 \\ 0 & 0 & 0 & 0 & \phi_{j1} & \phi_{j2} & -\hat{K_{j7}} \end{bmatrix},
$$

and,

$$
\hat{M_j} = [(S^* - S) + (1-\varepsilon_{jf})(V_f^* - V_f) + (1-\varepsilon_{jb})(V_b^* - V_b)] \begin{bmatrix} 0 & \beta_{jp} & \beta_{ja} & \beta_{jq} & \beta_{j1} & \beta_{j2} & \beta_{jh} \\ 0 & 0 & 0 & 0 & 0 & 0 & 0 \\ 0 & 0 & 0 & 0 & 0 & 0 & 0 \\ 0 & 0 & 0 & 0 & 0 & 0 & 0 \\ 0 & 0 & 0 & 0 & 0 & 0 & 0 \\ 0 & 0 & 0 & 0 & 0 & 0 & 0 \\ 0 & 0 & 0 & 0 & 0 & 0 & 0 \end{bmatrix}. \qquad \text{(S.2)}
$$

Since $S(t) \leq S^*$, $V_f(t) \leq V_f^*$ and $V_b(t) \leq V_b^*$ for all $t > 0$ in $\Omega_*$, it follows that the matrix $\hat{M_j}$, defined in (S.2), is non-negative. Hence, the equation (S.1) can be re-written in terms of the following inequality:

$$
\frac{d}{dt}\begin{bmatrix} E_j(t) \\ P_j(t) \\ A_j(t) \\ Q_j(t) \\ I_{j1}(t) \\ I_{j2}(t) \\ H_j(t) \end{bmatrix} \leq (\hat{F}_j - \hat{V}_j)\begin{bmatrix} E_j(t) \\ P_j(t) \\ A_j(t) \\ Q_j(t) \\ I_{j1}(t) \\ I_{j2}(t) \\ H_j(t) \end{bmatrix}. \qquad \text{(S.3)}
$$

It should be recalled from Theorem 3.1 that if $\hat{\mathbb{R}}_c < 1$, then all eigenvalues of the next generation matrices $\hat{F}_j\hat{V}_j^{-1}$ are negative (which is equivalent to $\hat{F}_j - \hat{V}_j$ been stable matrices [15]). Thus, it can be concluded that the linearized differential inequality system (S.3) is stable whenever $\hat{\mathbb{R}}_c < 1$. Hence, it follows from this analysis that (for this linear system of ordinary differential equations):

$$
(E_j(t), P_j(t), A_j(t), Q_j(t), I_{j1}(t), I_{j2}(t), H_j(t)) \to (0,0,0,0,0,0,0), \text{ as } t \to \infty.
$$

Substituting $E_j(t) = P_j(t) = A_j(t) = Q_j(t) = I_{j1}(t) = I_{j2}(t) = H_j(t) = 0$ into the differential equations for the rate of change of the $R_j(t)$, $S(t), V_f(t)$ and $V_b(t)$ compartments shows that:

$$
R_j(t) \to 0 \text{ and } S(t) \to S^*, V_f(t) \to V_f^*, V_b(t) \to V_b^* \text{ as } t \to \infty.
$$

Thus, the DFE (given in Equation (3.1) with $\omega_{vf} = 0$) for the special case of the model (2.8) (with $\delta_{j1} = \delta_{j2} = \delta_{jh} = 0$ and $\omega_{vf} = \omega_{dr} = \omega_{or} = 0$) is globally-asymptotically stable in $\Omega_*$ whenever $\hat{\mathbb{R}}_c < 1$. □

### S4 Assessing the Combined Impact of Vaccination and Mask Usage

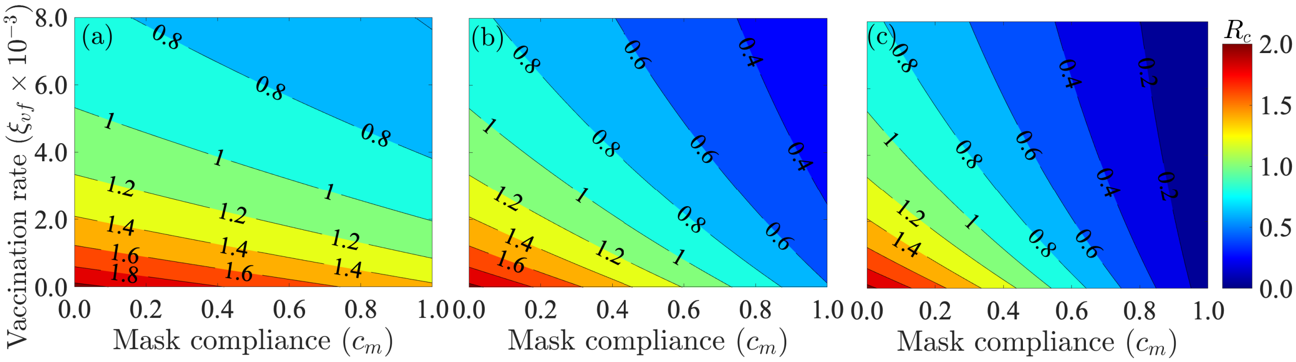


Figure S1: Contour plot of the control reproduction number of the model (2.8) as a function of the vaccine rate coverage ($\xi_{vf}$) and additional mask coverage ($c_m$) for additional (a) cloth mask coverage (i.e., $\varepsilon_m = 0.30$), (b) surgical mask coverage (i.e., $\varepsilon_m = 0.70$), and (c) N95 mask coverage (i.e., $\varepsilon_m = 0.95$). The other parameters used are given in Tables S3 and Table S4.

### S5 Assessing the Impact of Vaccination and Waning of Vaccine-derived Immunity

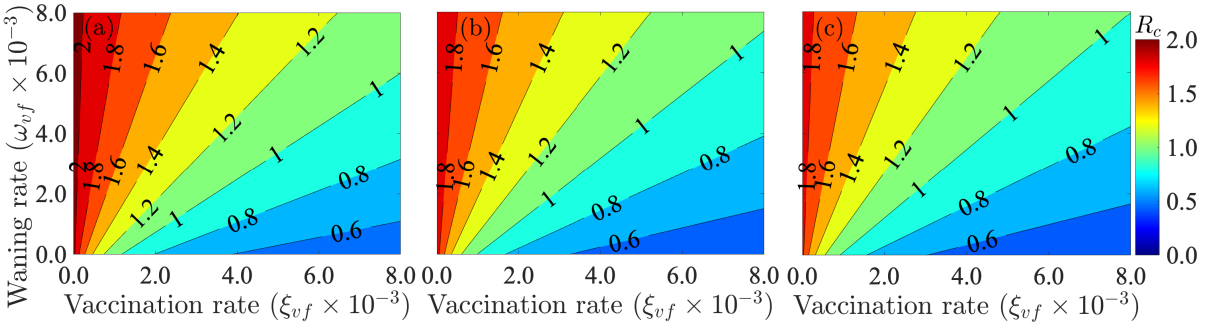


Figure S2: Contour plot of the control reproduction number of the model (2.8) as a function vaccine-derived immunity waning rate for fully vaccinated individuals ($\omega_{vf}$ per day) and the vaccination rate ($\xi_{vf}$ per day) when there is (a) no additional mask coverage beyond the baseline coverage, (b) a 10% increase in surgical mask compliance, (c) a 10% increase in N95 mask compliance. The other parameters used are given in Tables S3 and Table S4.

#### S5.1 Assessing the Impact of Detecting Exposed, Presymptomatic, and Asymptomatic Cases

The model (2.8) is further simulated to assess the impact of detection of exposed, presymptomatic and asymptomatic infectious humans (through testing) and the impact of the length of time it takes for vaccine-derived immunity to wane completely. For the first set of simulations, the detection rate of exposed, presymptomatic and asymptomatic infectious individuals with the Delta or the omicron variant, which are assumed to be the same in this study (i.e., $\rho_d = \rho_o$) is scaled by 25, 50, and 75. The simulation results obtained, depicted in Figure S3 (a) and (c), show an increase in the confirmed daily (Figure S3 (a)) and cumulative (Figure S3 (c)) cases of COVID-19 in the United States. with increasing detection of exposed, presymptomatic and asymptomatic infectious individuals. For example, if the detection rate is 25 times the baseline value, there is a 4% increase in the peak number of confirmed cases (magenta curve in Figure S3(a)). However, if the detection rate increases to 75 times the baseline value, then there is a 9% increase in the peak number of confirmed cases (green curve in Figure S3(a)). Similar increases in cumulative cases are recorded as the detection rate of exposed, presymptomatic and asymptomatic infectious individuals increases (Figure S3 (c)).

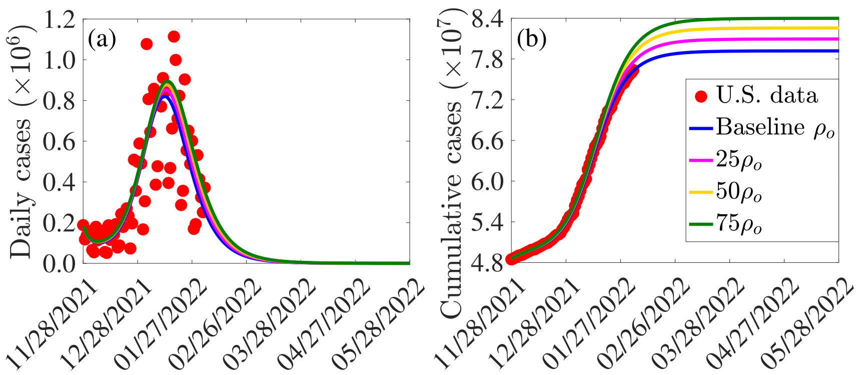


Figure S3: Simulations of the model (2.8) depicting the impact of detecting exposed, presymptomatic, and asymptomatic infectious individuals on the (a) the new daily and (b) cumulative COVID-19 cases in the US. The other parameter values used for the simulations are presented in Tables S3 and S4.
